# Design and selection of drug properties to increase the public health impact of next-generation seasonal malaria chemoprevention: a modelling study

**DOI:** 10.1101/2023.10.13.23292651

**Authors:** Lydia Braunack-Mayer, Josephine Malinga, Thiery Masserey, Narimane Nekkab, Swapnoleena Sen, David Schellenberg, André-Marie Tchouatieu, Sherrie L Kelly, Melissa A Penny

**Author notes:** These authors contributed equally (Josephine Malinga, Thiery Masserey, Narimane Nekkab). Correspondence to: Prof Melissa A Penny, Swiss Tropical and Public Health Institute Kreuzstrasse 2, 4123 Allschwil Switzerland.

## Abstract

**Background:** Seasonal malaria chemoprevention (SMC) is recommended for disease control in settings with moderate to high *Plasmodium falciparum* transmission and currently depends on administration of sulfadoxine-pyrimethamine with amodiaquine. However, poor regimenadherence and the increasedfrequencyof sulfadoxine-pyrimethamine resistant parasite mutations may threaten SMC’s effectiveness. We need guidance to de-risk the development of drug compounds for malaria prevention.

**Methods:** We combined an individual-based malaria transmission model that has explicit parasite growth with drug pharmacokinetic/pharmacodynamic models. We modelled SMC drug attributes for several possible modes-of-action, linked to their potential public health impact. Global sensitivity analyses identified trade-offs between drug elimination half-life, maximum killing effect, and SMC coverage, and optimisation identified minimum requirements to maximise malaria burden reductions.

**Findings:** Model predictions show that preventing infection for the entire period between SMC cycles is more important than drug curative efficacy for clinical disease effectiveness outcomes, but similarly important for impact on prevalence. When four SMC cycles are deployed to children under five years with high levels of coverage (69% of children receiving all cycles), drug candidates require a duration of protection half-life of >23 days (elimination half-life >10 days) to achieve >75% clinical incidence and severe disease reductions (measured over the intervention period in the target population, compared with no intervention across a range of modelled scenarios). High coverage is critical to achieve these targets, requiring >60% of children received all SMC cycles and >90% of children at least one cycle regardless of the drug’s duration.

**Interpretation:** While efficacy is crucial for malaria prevalence reductions, chemoprevention development should select drug candidates for their duration of protection to maximise burden reductions, with the duration half-life determiningcycle timing. Explicitlydesigning or selectingdrug properties to increase communityuptake is paramount.

**Funding:** Bill & Melinda Gates Foundation and the Swiss National Science Foundation.

## Background

Seasonal malaria chemoprevention (SMC) is recommended for *Plasmodium falciparum* malaria control in areas of seasonal transmission to reduce disease burden among children belonging to age groups at high risk of severe malaria.^1^ SMC has so far depended on a complete treatment course of one dose of sulfadoxine-pyrimethamine (SP) plus three daily doses of amodiaquine (AQ), delivered at monthly intervals. Through these multiple deployments of an antimalarial drug, SMC maintains therapeutic drug concentrations throughout the high-risk period. This intervention has proven effective when delivered as part of routine malaria control, with a mean 88·2% (95% CI 78·7−93·4%) reductioninthe incidence of clinical malariaamong childrenunder five years within 28 days of administration following each cycle of SMC, and a 42·4% (95% CI 5·9−64·7%) and 56·6% (95% CI 28·9−73·5%) mean reduction in malaria-related deaths in Burkina Faso and in The Gambia, respectively.^2^

The spread of partial *P. falciparum* resistance to SP may eventually threaten SMC’s effectiveness. Increasing frequency of the quintuple mutation associated with SP resistance (a triple mutation in *pfdhfr* plus *pfdhps*-437Gly and *pfdhps*-540Glu) has been observed in the sub-Sahel region.^2^ Although, the overall frequency of the quintuple mutation and the frequency of mutations conferring partial resistance to AQ (*Pfcrt-CIVET* + *pfmdr1-86 Tyr* + *184 Tyr*) remains low.^3^ SP resistance has been observed to reduce the duration of protection afforded by the drug in infancy and pregnancy,^4,5^ and may threaten SMC’s ability to provide individual protection and population-level effectiveness.^6–8^ SMC appears to be maintaining its effectiveness as a public health control measure.^9,10^ However, this may not continue if the frequency of resistant mutations increases. More evidence is needed to assess this risk.

To address the need for new SMC drugs, a clinical pipeline is being developed to repurpose existing drugs and to make new drugs available. Emerging guidelines have characterised target product profiles (TPPs) for chemoprevention interventions^11^ and, for the first time, funders and drug developers have targeted the development of candidates specifically for chemoprevention. Medicines for Malaria Venture (MMV)’s TPP-2 for chemoprevention^12^ underpins the research and development organisation’s candidate pipeline, such as for MMV371 and ganaplacide. New SMC combination drug candidates have also been identified from a set of existing drugs used to treat clinical malaria or for prevention in travelers, such as atovaquone-proguanil (ATV-PG),^13^ piperaquine (PPQ), and pyronaridine (PYN).^11^

Yet, in the early stages of product development, we do not know which drug characteristics will be the most important for SMC’s continued effectiveness. Emerging guidelines are benchmarking desired properties for drug candidates based on the properties and protection afforded by SP-AQ,^14^ without yet explicitly defining how these drugs can address the shortcomings of this standard-of-care. In particular, candidate selection will involve balancing desirable pharmacokinetics (PK) and pharmacodynamics (PD) properties with SMC’s delivery characteristics. However, to the best of our knowledge, no study has considered how a drug candidate’s duration of protection, protective efficacy, and implementation coverage can be balanced to increase SMC’s effectiveness. By duration of protection, we refer to the length of time for which an individual is protectedagainst infectionafter receiving a drug. By protective efficacy, we refer to a drug’s within-host efficacy against one or more stages of the parasite lifecycle. This term is distinct from effectiveness, which refers to the clinical cases averted by deploying a drug in a population. By implementation coverage, we refer to the proportion of SMC’s target population with access to the intervention.

SMC’s likely impact with SP-AQ beyond that observed in clinical trials has been shown in several observational studies and further explored in various studies with models built on a range of assumptions around the intervention’s action on *P. falciparum*’s blood or liver stages.^15–18^ However, the mechanism-of-action for existing, alternative, and new SMC drugs has yet to be fully understood. Furthermore, no single existing study has integrated uncertainty around the unknown pharmacological properties of next-generation drugs. Quantitative evidence that better accounts for these complexities is needed to define minimum product criteria for next - generation SMC and to increase success rates for SMC drug development.

To address these knowledge gaps, we used mathematical and statistical modelling approaches to quantify minimum criteria for characterising the next-generation of SMC drugs. We generated this evidence through engagement with malaria chemoprevention experts and through use of an individual-based malaria transmission model to analyse likely public health outcomes of three probable mechanisms of action for a next generation of SMC drugs. Through this evidence, we aim to provide funders and drug developers with guidance for the early prioritisation of new and alternative SMC drugs and their target product profile documents.

## Methods

### Expert consultation

This study was grounded in findings from consultations with chemoprevention drug and guideline developers.^19^ Experts identified the need to, first, quantify the impact of trade-offs between intervention characteristics (efficacy, duration of protection, coverage, and dosing) on public health outcomes and, second, set preferred ranges around these characteristics towards achieving health targets for next-generation SMC. These discussions, as well as ongoing feedback from malaria chemoprevention experts, shaped the use cases we considered and underpinned our approach to estimating the public health impact of SMC product candidates.

### Malaria transmission model

We applied an established individual-based malaria transmission model, OpenMalaria (https://github.com/SwissTPH/openmalaria/wiki), to predict the impact of next-generation SMC products on population-level outcomes. The model and parameterisations used in this work have been fully described previously^20–22^ and are summarised in appendix 1.1. OpenMalaria consists of different model components representing the chain of processes from the mosquito lifecycle to malaria infection, treatment, and immunity acquisition of a human host, and captures differences in consequences of immunity on care seeking and clinical disease.^23^ The model variant^20^ used for this study includes a mechanistic within-host model component for the parasite lifecycle in humans, which describes the time-course of asexual *P. falciparum* parasitaemia following a single inoculation. Transmission from infected humans to mosquitoes depends on this asexual parasite density, with gametocyte densities following between ten and 20 days later.^22–24^ This model also incorporates PK/PD models for a comprehensive set of intervention dynamic characteristics.

The malaria transmission model was used to simulate a range of transmission and health system scenarios. Each scenario simulated a unique combination of setting characteristics (appendix table A1.2), including: two levels of health system access to first-line malaria treatment (10% and 50%); two malaria transmission seasonality profiles (70% of cases occurring within a three- and five-month period, respectively), and; a range of transmission intensities from low (8%) to high (39%) baseline annual prevalence rate of *P. falciparum* rapid diagnostic test (RDT) detectable infections in children two to ten years of age (*Pf*PR_2-10_) under a no SMC counterfactual.

### Intervention dynamics and outcome measures

Using models to infer a drug-based intervention’s likely impact relies on an accurate understanding of where and how the drug acts. However, for many SMC candidates, clinical evidence of drug activity and protective efficacy is not yet available. For this reason, we describe an SMC candidate’s likely effect by using PK/PD models of three plausible mechanisms of action: blood stage activity only; SMC with dominant blood stage activity and initial, complete liver stage clearance, and; SMC with dominant liver stage activity and initial, complete blood stage clearance. For all three mechanisms, we represent the uncertainty inherent in modelling drugs whose PK/PD properties are not yet fully known by modelling a range of plausible drug properties and deployment coverage characteristics (table 1). Intervention modelling assumptions are fully described in appendix 1.3.

**Table 1:**
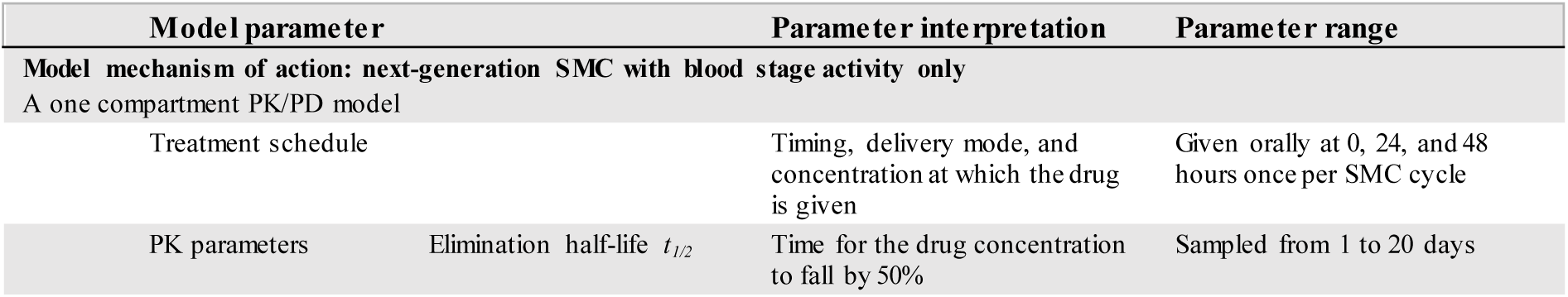
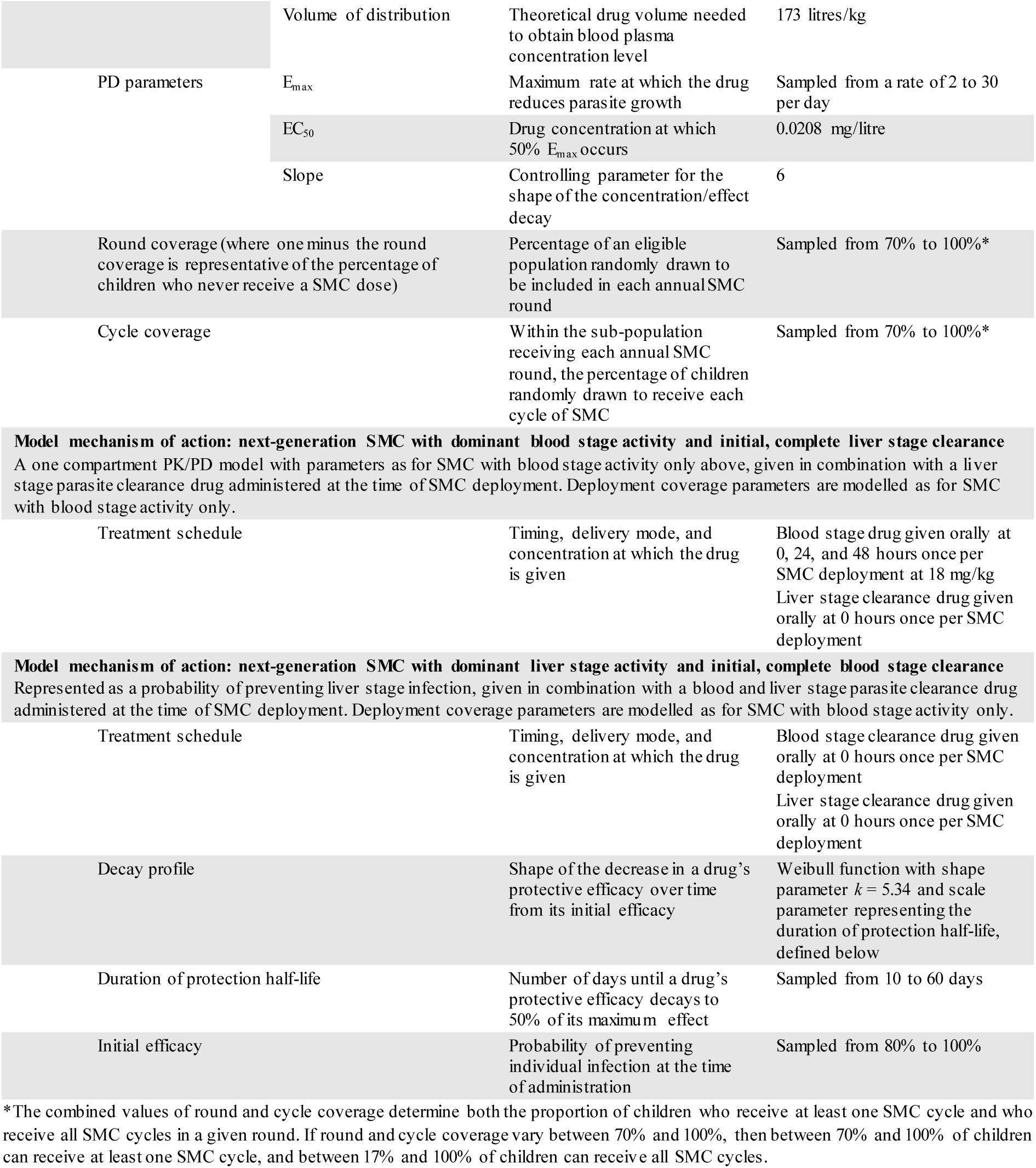
Summary of model characteristics and parameter ranges for each of the three mechanisms for an SMC candidate’s effect. We modelled three plausible mechanisms of action that describe the drug’s or drug combinations’ activity against blood stage or liver stage parasitaemia. Intervention modelling assumptions are fully described in appendix 1.3. Our first approach to modelling next-generation SMC captured the intervention’s effect on reducing the growth rate of asexual blood stage parasitaemia. We used a one-compartment PK/PD model to describe the drug’s killing effect over time,^25^ referred to as SMC with blood stage activity only. For simplicity, given that both PK/PD properties for next-generation SMC and PD properties for SP for are still unknown (unpublished; Masserey, Braunack-Mayer, and colleagues), we assumed piperaquine-like behavior and dosing.^26^ Second, we considered a candidate with both blood and liver stage activity by adding a model component that clears liver stage parasitaemia at the time of drug administration. This second model is referred to as SMC with dominant blood stage activity and initial, complete liver stage clearance. Third, we considered candidates with predominantly liver stage activity by deploying a previously calibrated model for SP-AQ with protection against new infections that decays over time and with blood and liver stage parasite clearance at the time of drug administration.^27^ This model is referred to as SMC with dominant liver stage activity and initial, complete blood stage clearance. Unless specified, model properties and parameter ranges are the same across approaches.

This complex modelling approach was not intended to be used to compare the impact of drug activity on SMC’s effectiveness, but rather was necessary to capture a broad spectrum of drug properties across the entire chemoprevention candidate development pipeline. By doing so, we provide ranges to inform an understanding of candidate impact.

We explored the importance of next-generation SMC drugs characteristics for different deployments by simulating outcomes for three, four, or five cycles of SMC delivered to children aged three to 59 months. In line with WHO terminology, we use ‘cycle’ to refer to the monthly administrations of SMC in a given year and ‘round’ to refer to each annual deployment of an SMC program. For deployments with three and four cycles, the first cycle was deployed in the month prior to the seasonal peak. For five cycles, the first cycle began two months prior to the seasonal peak (appendix figure A1.2). We also modelled SMC for a broader target population – children aged three to 119 months – with results presented in appendix section two. Primary outcome measures were the reduction in clinical incidence, prevalence, and severe disease incidence in the target populations over the three-, four-, or five-month intervention periods five years after SMC’s first deployment. Reductions were calculated relative to a no SMC counterfactual measured in the same period and target populations (children aged three to 59 months) from the year prior to SMC’s introduction. All outcome measures and deployment characteristics are fully described in appendix table A1.2.

### Standard-of-care parameters

Clinical evidence of drug activity, duration, and protective efficacy is not yet available for many SMC candidates. For this reason, in the absence of data for model validation, we assessed the ability of our three drug models to accurately represent the dynamics of SP-AQ, SMC’s standard-of-care. We performed this analysis by replicating protective efficacy outcomes from a randomised non-inferiority trial of dihydroartemisinin-piperaquine (DHA-PPQ) to SP-AQ,^26^ following the approach described by Burgert and colleagues.^27^ SP-AQ’s protective efficacy against clinical case incidence was extracted from Zongo and colleagues.^26^ For each SMC modelling approach (blood stage only, dominant blood stage, and dominant liver stage), we used OpenMalaria to perform in-silico clinical trials for a wide range of model parameter values. We then identified the range of parameter values that produced SP-AQ-like results by minimising the residual sum of squares (RSS) between simulated trial outcomes and SP-AQ’s protective efficacy. This approach is fully described in appendix 1.3.

### Simulation and statistical analysis

Our analysis then applied a mathematical framework for predicting determinants of intervention impact and defining minimum drug profile criteria. This framework has been previously described,^28^ and was demonstrated in a proof-of-concept study^27^ (as summarised in appendix 1.4). In brief, we used the individual-based malaria transmission model to simulate scenarios over a wide range of input values for intervention coverage and drug initial efficacy and duration of protection. Heteroskedastic Gaussian Process regression models were then trained to capture the relationship between intervention inputs and public health outcomes. These machine learning models permitted us to explore a large parameter space with fewer calls to the computationally intensive simulation model. Using these model emulators, we performed a global sensitivity analysis using the Sobol-Janssen method^29^ to identify synergisms between intervention characteristics. We also used optimisation methods to identify minimum drug criteria towards achieving a target outcome.

## Results

For SMC with blood stage only and dominant blood stage activity, we found that an elimination half-life of 4·74 to 8·81 and 5·12 to 8·81 days, respectively, produces SP-AQ-like behavior, a range comparable to published studies of SP’s pharmacokinetics.^30^ For SMC with dominant liver stage activity, SP-AQ’s protective efficacy was represented by a drug with a duration of protection half-life (representing the time until drug’s protective efficacy reaches 50% of its initial value, a different metric from elimination half-life) of 23·78 to 29·83 days (figure 1 and table 2).

**Figure 1:**
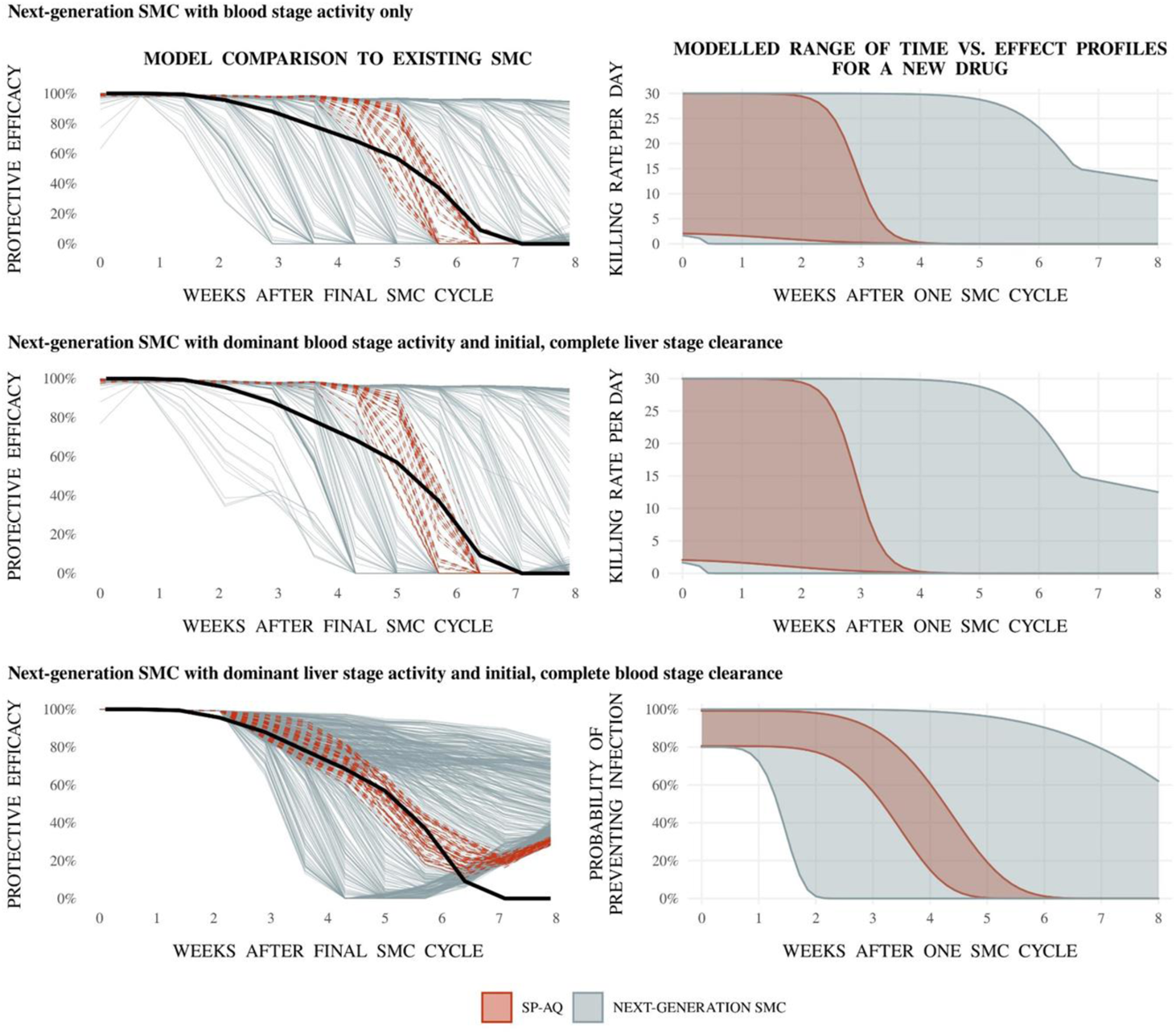
Model comparison to SP-AQ’s protective efficacy profile as reported in randomised non-inferiority trial of SMC conducted in Burkina Faso^26^ for each of three SMC modelling approaches. **Left panels** show the protective efficacy of in-silico clinical trial outcomes for each simulated next-generation SMC profile. Grey curves indicate the model-estimated protective efficacy against clinical case incidence of each of 500 simulated SMC drug profiles. Solid black curves indicate SP-AQ’s protective efficacy against clinical case incidence as extracted from figure 3 of Zongo and colleagues.^26^ Red dashed curves indicate a selection of the 500 simulated SMC drug profiles with a similar model-estimated protective efficacy to SP-AQ. This selection was assessed by calculating the residual sum of squares (RSS) between simulated trial outcomes and SP-AQ’s protective efficacy and by identifying the simulated profiles with a RSS within 0·1 standard deviations of the minimum. **Right panels** show the full parameter space of modelled time-effect relationships for a new drug (grey shaded regions). The red shaded regions indicates the space of parameter profiles with a similar model-estimated protective efficacy to SP-AQ, as indicated in the left panels.

**Table 2:**
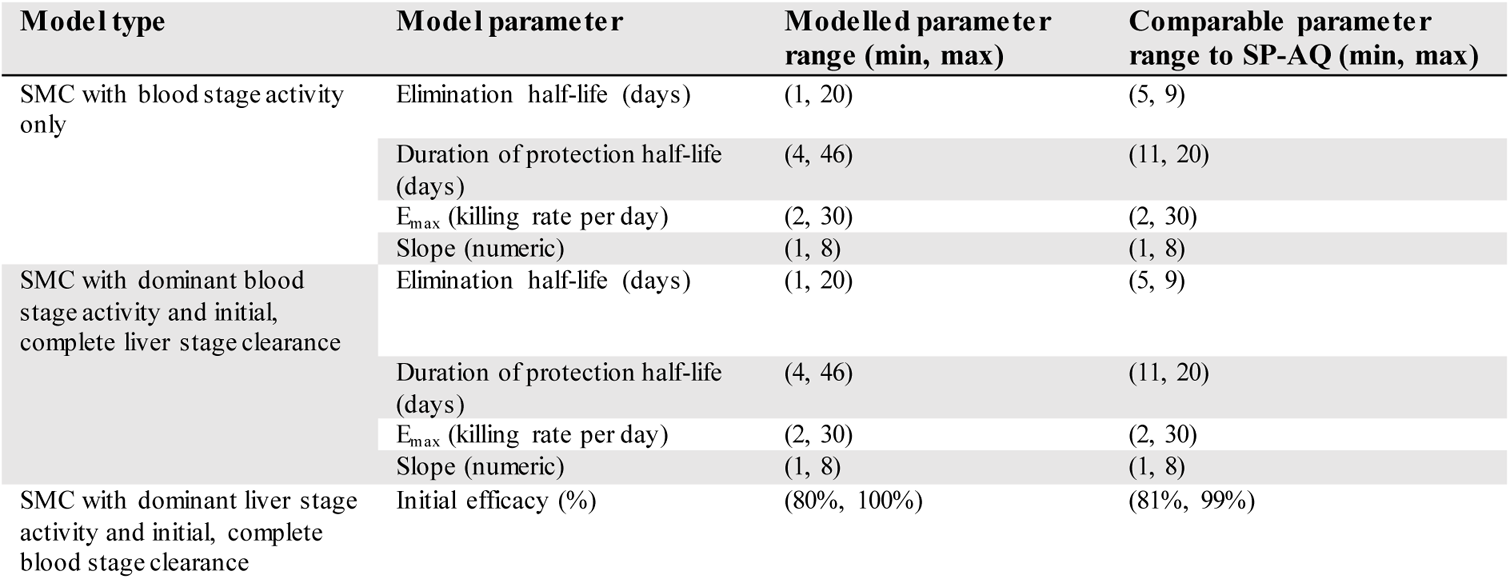
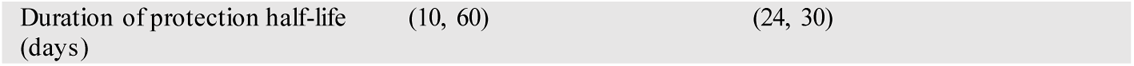
Comparable range of SMC model parameters to SP-AQ. For each of the three modelling approaches, minimum and maximum values for the range of modelled parameters with a similar protective efficacy to SP-AQ (RSS for in-silico protective efficacy in comparison to SP-AQ’s protective efficacy falling within 0·1 standard deviations of the minimum). SP-AQ’s protective efficacy against clinical case incidence was extracted from figure 3 of Zongo and collagues.^26^

For all three drug models and across the range of intervention properties evaluated in this study, our analysis confirmed that an SMC candidate’s duration of protection is critical for minimising malaria morbidity. For SMC with blood stage activity only, drug elimination half-life was the most important driver of impact on clinical incidence, prevalence, and severe disease reduction across the majority of modelled scenarios (appendix figures A2.1, A2.3, and A2.6). Elimination half-life was the most or second-most important driver for SMC with dominant blood stage activity and initial, complete liver stage clearance (figure 2, appendix figures A2.4 and A2.7). For example, when SMC with dominant blood stage activity was deployed to children aged three to 59 months, this characteristic explained up to 54% of variation across all outcome measures and across all modelled scenarios. Duration for SMC with dominant liver stage activity had a lesser importance (appendix figures A2.2, A2.5, and A2.8), driven by the model assumption of blood and liver stage parasite clearance at the time of administration.

**Figure 2:**
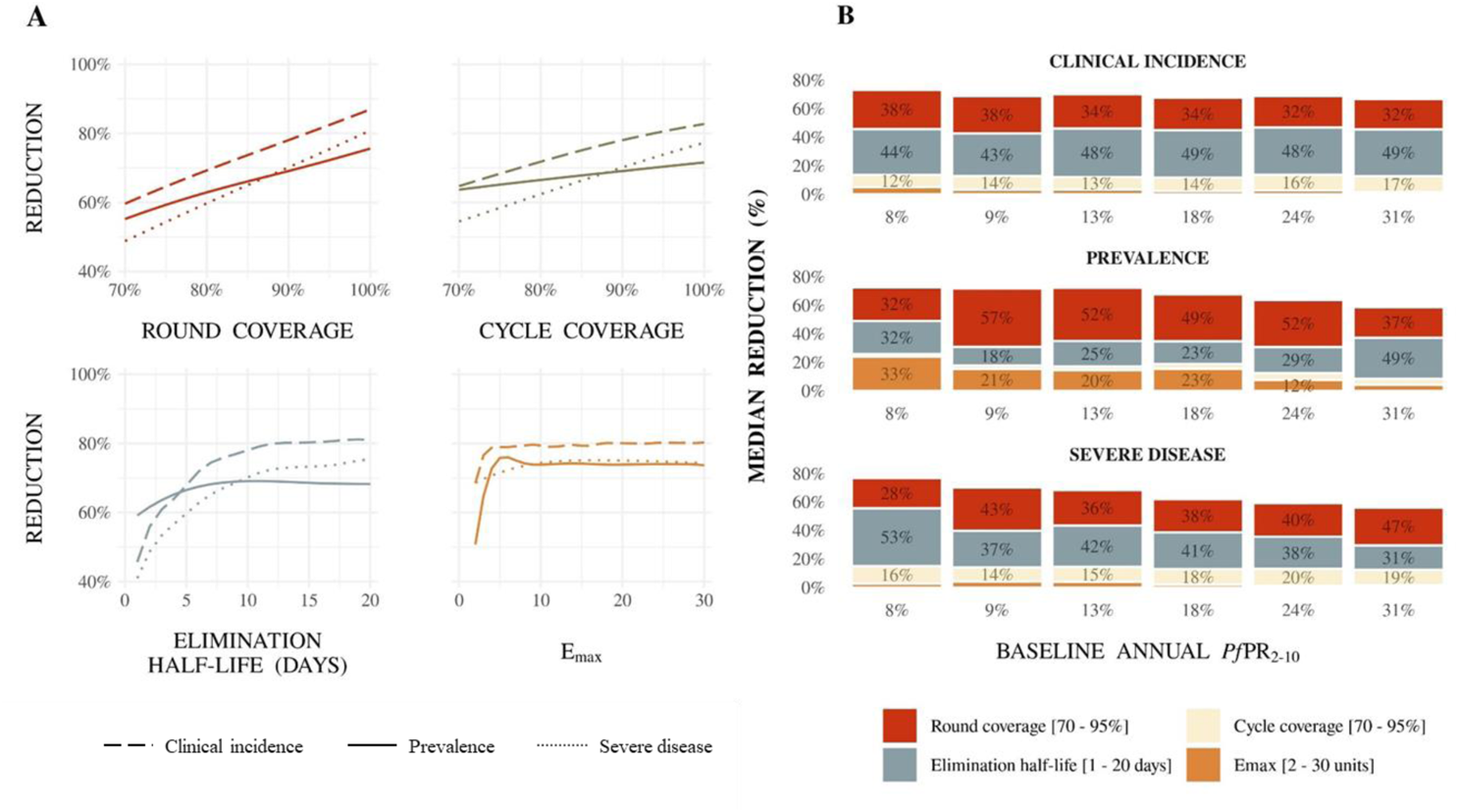
For SMC with dominant blood stage activity and initial, complete liver stage clearance, (A) illustrative Gaussian Process regression emulator predictions for the relationship between SMC properties and clinical incidence, prevalence, and severe disease reductions, and (B) drivers of impact on predicted clinical incidence, prevalence, and severe disease reduction for SMC in children three to 59 months of age, compared with a no intervention counterfactual. (A) Results show a scenario with 50% access to first-line treatment over 14 days and a five-month seasonal profile with baseline annual *Pf*PR_2-10_ of 18% when SMC was deployed in four monthly cycles to children aged three to 59 months in a given year. Each panel shows the predicted reduction when all performance characteristics but the parameter of interest were held constant at: 90% round coverage, 90% cycle coverage, an elimination half-life of 10 days, and a maximum parasite killing rate of 3.45 units. Alternative profiles for these constant paramet ers are shown in appendix figure A2.9. (B) Bars indicate the total Sobol effect indices for key model parameters. Indices can be interpreted as the proportion of va riation in the outcome attributable to a given change in each variable, along with its interactions with other variables. Bar heights indicate the median expected reduction across the modelled parameter range. Results are shown across prevalence settings (horizontal axis, baseline annual *Pf*PR_2-10_) for a scenario of 50% access to first-line treatment within 14 days with a five-month seasonal profile, where SMC was deployed in four monthly intervals in a given year.

Our modelling suggests that an intervention’s initial killing effect was important to reduce malaria prevalence in the target population. In our models, initial killing can be thought of as capturing the impact of adherence to a drug’s dosing schedule or of the drug’s initial treatment efficacy. In the setting shown in figure 2, the drug’s rate of blood stage parasite clearance accounted for up to 33% of variation in prevalence reduction across baseline transmission settings. The importance of a drug’s killing effect suggested that prevalence reduction was driven by both the effectiveness with which a drug cleared blood stage parasites, as well as the duration over which the drug prevented new infections from taking hold. Drug elimination half-life remains more important than the killing effect for impact on clinical disease. Our individual-based model’s ability to capture differences in immunity means that clinical incidence and prevalence endpoints provide a mechanism to translate across different settings and, thus, can facilitate the benchmarking of endpoints between different clinical trials and SMC candidates.

In addition to a drug’s elimination half-life and blood stage activity, SMC coverage was a critical determinant of public health impact. Our sensitivity analysis of results for SMC with dominant blood stage activity and initial, complete liver stage clearance indicated that, across the range of modelled scenarios, between 35% and 80% of variation in clinical incidence, prevalence, and severe disease reductioncouldbe attributed to the combinedimpact of SMC’s round coverage (percentage of children with access to an SMC round, meaning to the three, four, or five cycles of an annual SMC campaign) and cycle coverage (percentage of children within those with access to a round who also receive each SMC cycle) (figure 2B). As both round and cycle coverage increased, SMC was predicted to reduce the burden of clinical cases in children who receive the intervention (figure 2A). Coverage was similarly critical for the burden reduction of SMC modelled as dominant liver stage activity and initial, complete blood stage clearance but was less important than the elimination half-life when SMC was modelled with blood stage activity only, driven by the fact that drug duration makes up for the lack of complete liver stage parasite clearance at the time of drug administration in this model (appendix figures A2.1 and A2.2).

SMC coverage and elimination half-life were the key impact drivers for next-generation SMC across seasonal profiles (three- and five-month) and across levels of access (10% and 50%) to first-line curative treatment for malaria (appendix figures A2.6, A2.7, and A2.8). Similar results were observed when SMC was deployed to children three to 119 months (appendix figures A2.3, A2.4, and A2.5). We also observed evidence of very low indirect effects of SMC on children up to 119 months not receiving SMC, particularly for drug candidates with high coverage and long elimination half-life (appendix figure A2.10).

Our results highlight a potential to trade a reduced dosing schedule and, hence, potentially shorter elimination half-life in favour of facilitating higher SMC coverage for impact on clinical incidence and severe disease. In moderate transmissionintensitieswith 18% *Pf*PR_2-10_, where SMC was deployed with four monthly cycles annually to children aged three to 59 months, an intervention with an elimination half-life of ten days and E_max_ of ten was predicted to achieve a 74% reduction in clinical incidence (95% prediction interval 67%−80%) when both round and cycle coverage were 85% (figure 3). Deploying the same intervention with 95% round coverage led to an increase in the expected clinical incidence reduction to 83% (95% prediction interval 79%−87%). Similar trade-offs were apparent for SMC with blood stage only activity (appendix figure A2.11) and dominant liver-stage activity and initial, complete blood stage clearance (appendix figure A2.12).

**Figure 3:**
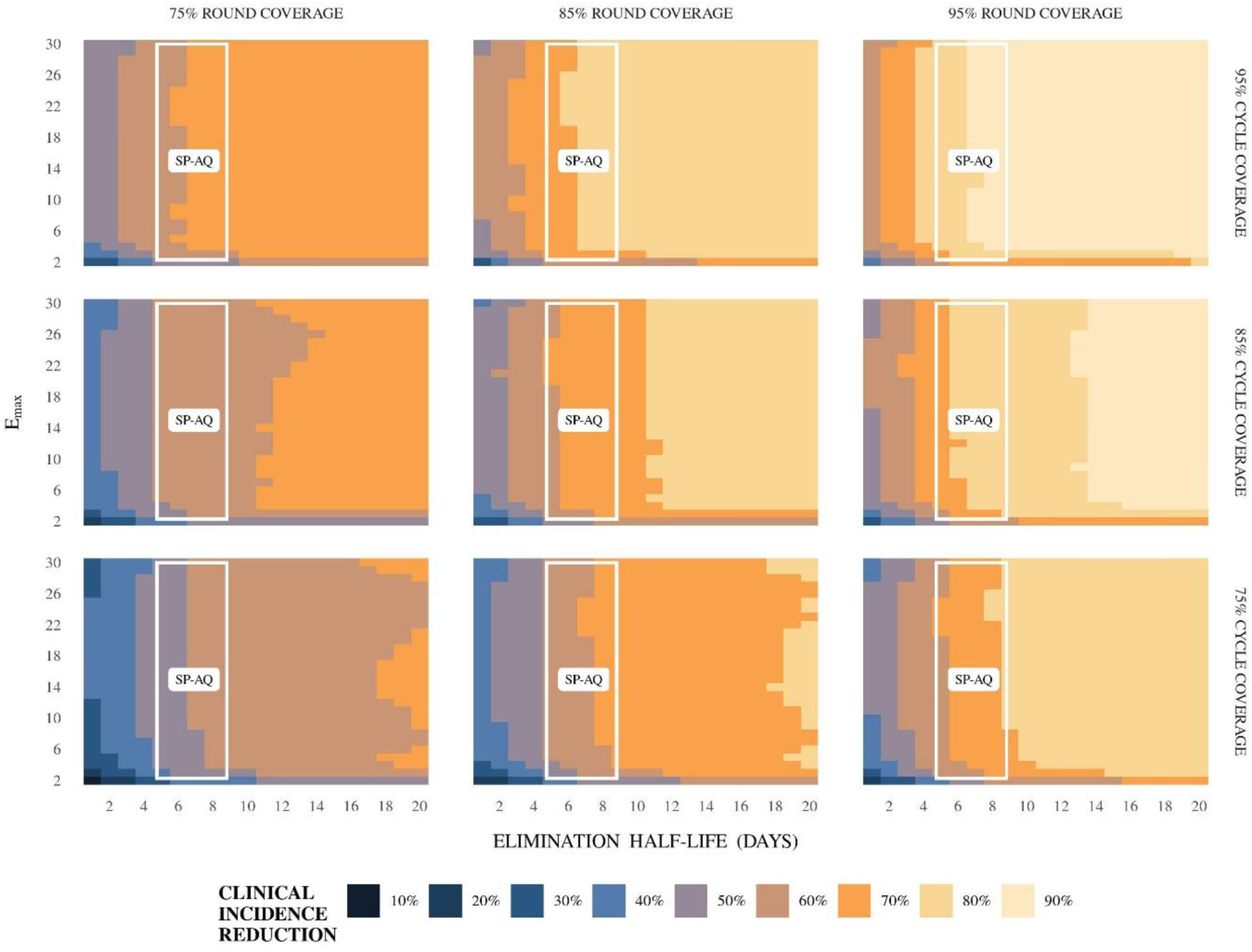
For SMC with dominant blood stage activity and initial, complete liver stage clearance, impact of a change in coverage on the predicted relative reduction in clinical incidence (measured over a four-month intervention period) following SMC compared with a no intervention counterfactual. Each square in the grid indicates the predicted reduction (rounded to the nearest 10%) if an intervention with the given elimination half-life (horizontal axis) and maximum parasite killing rate (vertical axis) were deployed, assuming a slope of six. Variation in this figure is driven by the combined impact of stochastic uncertainty and emulator prediction error. Results are shown for children aged three and 59 months for a five-month seasonal profile with an baseline annual *Pf*PR_2-10_ of 18%, where access to first line treatment was 50% within 14 days and where SMC was deployed four times at monthly intervals in a given year surrounding peak seasonality. Each panel represents results for a different level of SMC round coverage (75%, 85%, and 95%) and cycle coverage (75%, 85%, and 95%). The white region indicates the space of parameter profiles with a similar protective efficacy to SP-AQ (residual sum of squares for in-silico protective efficacy in comparison to SP-AQ’s protective efficacy falling within 0·1 standard deviations of the minimum, as in figure 1).

Together these results indicate that, for greater public health impact, SMC requires an intervention with an extended duration of protection between cycles, and with high coverage. Following an optimisation procedure, we identifiedminimum requirements for these keyinterventioncharacteristicsacrossmodelledscenarios for SMC with dominant blood stage activity and initial, complete liver stage clearance. For SMC’s coverage, our results indicate that over 60% of children should receive all cycles and over 90% at least one cycle of SMC to achieve targets of 75% clinical incidence and severe disease reduction across transmission settings (figure 4A). Extended protection between SMC cycles is also required. For example, for a scenario where SMC was delivered in four monthly cycles to children aged three to 59 months with high coverage (85% round coverage and 95% cycle coverage, translating to 69% of children receiving all SMC cycles), a duration of protection half-life of more than 23 days was required to achieve a 75% clinical incidence and severe disease reduction (figure 4B). This duration half-life is longer than the duration half-life we estimated for SP-AQ in our model validation exercise, suggesting scope to further optimise SMC with SP-AQ by shortening the time between cycles.

**Figure 4:**
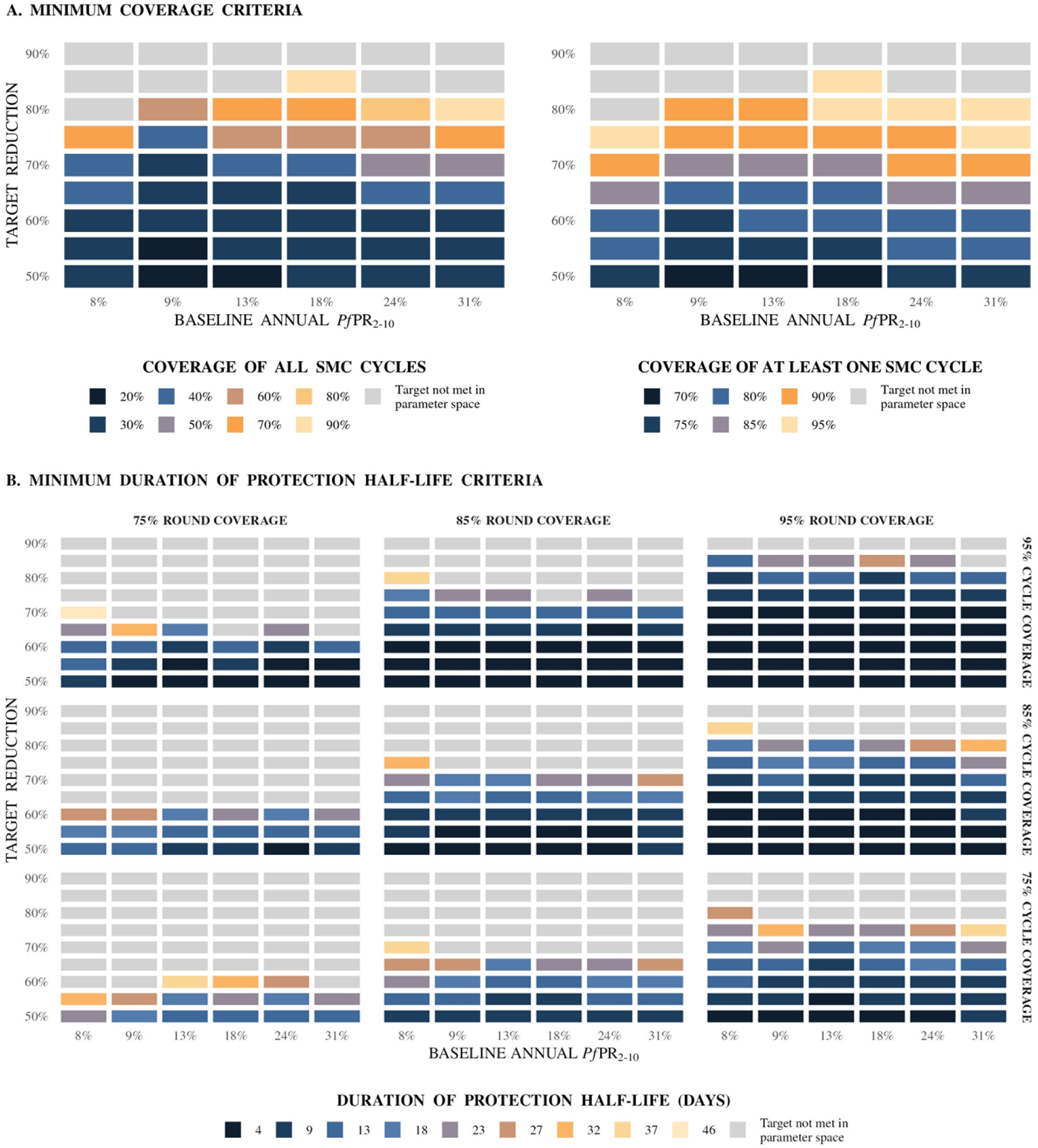
Summary of predicted minimum parameter values for SMC with dominant blood stage activity and initial, complete liver stage clearance to achieve target reductions in clinical incidence and severe disease. (A) Summary of the predicted minimum coverage criteria for SMC with dominant blood stage activity and initial, complete liver stage clearance towards achieving target reductions in clinical incidence and severe disease across SMC deployments, compared with a no intervention counterfactual. Results show estimated minimum values for SMC coverage characteristics whose 95% prediction interval was above the given target reduction for both clinical incidence and severe disease (vertical axis), aggregated across outcomes (clinical incidence and severe disease reductions measured across the intervention period), SMC deployments (three, four, and five monthly cycles of SMC in a given year) and levels for other model parameters (elimination half-life and E_max_). Results are shown for a five-month seasonal profile in a setting with 50% access to first-line treatment, where SMC was deployed to children three to 59 months old. (B) Summary of the predicted minimum duration of protection half-life criteria for SMC with dominant blood stage activity and initial, complete liver stage clearance towards achieving a target clinical incidence reduction for varying levels of SMC round and cy cle coverage, shown for a five-month seasonal profile in a setting with 50% access to first-line treatment, where SMC was deployed in four monthly cycles to children three to 59 months old. Results show the estimated minimum duration of protection half-life with a 95% prediction interval above the given target reduction in clinical incidence (vertical axis), compared with a no intervention counterfactual and measured across the four-month intervention period. Duration of protection half-life is defined as the number of days until a drug’s killing effect reaches 50% of its maximum effect. Minimum criteria were calculated across E_max_ levels.

## Discussion

We used an individual-based model of malaria transmission combined with multiple mechanisms of action and a broad range of drug properties, validated with clinical trial data for SP-AQ, to link individual-level drug characteristics to the likely population-level impact of new drugs for SMC. We identified which drug and intervention characteristics were the most important contributors to SMC’s impact on public health. We found that impact on clinical incidence was primarilydeterminedby SMC’s campaign coverage and by a drug’s duration of protection. More specifically, for spacing of four weeks between SMC cycles, a duration of protection longer than currently provided by SP-AQ was required to minimize clinical incidence and maximise prevalence reductions. If, however, we aimed to reduce prevalence, then high initial efficacy was also essential. Importantly, these findings quantitatively demonstrate that the ideal drug profile for malaria treatment is different from the ideal chemoprevention profile; the former requires high efficacy and does not require long duration of protection, while the latter can have lower efficacy but must have a long duration.

Our findings quantify the intuitive notion that a chemoprevention candidate’s duration of protection is essential for determining which candidates are likely to have the greatest effectiveness beyond clinical trials. Modelling three, four, and five cycles of SMC allowed us to evaluate how the ideal drug duration might change if SMC’s deployment does not cover the full malaria transmission season. Varying cycle timing highlighted that, to minimise clinical incidence and maximise prevalence reductions, the length of time between SMC cycles should be matched to the drug’s duration of protection. Furthermore, in our blood stage models, drug duration was varied through the candidate’s elimination half-life. In reality, a drug’s duration of protection will be determined by the interplay between PK and PD characteristics, including its EC_50_, minimal effective concentration, and effect decay profile. For drug combinations, duration of protection will be further influenced by the presence of drug-drug interactions. The precise balance of potency, PK, and host interactions required to maximise duration will be specific to each product, and these will need to be determined by drug developers for each chemoprevention candidate.

The importance of duration of protectionfor a chemopreventiondrug confirms the necessityof routine monitoring of chemoprevention efficacy against any infection endpoints, which should be measured in both early and late stage clinical studies of PK/PD properties, and when chemoprevention is routinely deployed.^31^ Ideally this would be supported through human challenge studies of drug activity against both blood and liver stage parasites, such as for the recently completed trial for cabamiquine.^32^ Natural infection phase two studies with highly sensitive PCR endpoints can also be used. Published PK/PD evidence from clinical trials is available for some combination SMC candidates, such as for ATV-PG with AQ.^13^ This particular combination showed unanticipated safety signals, reinforcing the need for rigorous assessment of repurposed drugs when used in combination. However, to the best of our knowledge, no published clinical trial has looked for synergies between PK and PD properties of combination SMC candidates ATV-PG with CQ, PPQ, or PYN, and, as a result, evidence for the impact of such synergies on an SMC candidate’s duration of protection is limited. At the time of writing, two ongoing trials (clinicaltrials.gov identifiers NCT03726593 and NCT05689047) are assessing efficacy outcomes for some of these chemoprevention combination candidates.

Regardless of a candidate’s duration, we need consensus across the malaria product development community on the importance of drug activity for chemoprevention’s impact to make informed drug selection decisions. Crucially, the clinical trial evidence described above is not available for SP-AQ, and our knowledge of SP’s action against liver stage parasitaemia is based on studies of pyrimethamine alone.^33^ In particular, recent evidence indicates that SP-AQ retains its effectiveness for chemoprevention within regions with moderate prevalence of the *dhfr/dhps* quadruple mutant associated with SP treatment failure.^9,10^ Yet, our lack of understanding of SP-AQ’s liver stage action means that the reasons for SP-AQ’s continued effectiveness remains unclear. Furthermore, it is posited that part of SP’s effectiveness in other chemoprevention programs, such as intermittent preventative therapy in pregnancy (IPTp), may be due to non-malarial pathways.^34^ Given the difficulty and cost of running clinical trials and observational studies, mathematical modelling has an important role to play in building consensus on drug activity. Further modelling studies could use scenario-based modelling with PK/PD models to develop hypotheses for SMC’s contribution to immunity acquisition, effectiveness against resistant parasites, and non-malarial pathways.

Interestingly, our results indicate that a chemoprevention drug’s parasite killing effect is of lower importance for its public health impact on averting clinical and severe disease than its elimination half-life. This confirms that the ideal chemoprevention drug profile is different to the ideal treatment profile, which must have high efficacy to clear parasites. A drug’s duration of protection is likely to have longer-term effects on immunity acquisition for SMC’s target population – children who may have low-level parasitaemia but who do not present with clinical malaria. A chemoprevention drug that prevents clinical symptoms but does not rapidly clear parasitaemia may allow children to build malaria immunity. Importantly, this may justify re-purposing antimalarial drugs that have failed to meet minimum treatment efficacy thresholds.

Repurposing an antimalarial drug that may not offer very high efficacy but has a long duration does, however, raise a number of considerations. First, we must consider that permitting low-grade parasitaemia may have a negative impact on a child’s health. Second, by deploying a chemoprevention product that does not rapidly clear parasitaemia, we may increase the risk of parasite resistance. This may, in turn, reduce the usable lifespan of a chemoprevention product. It may also require that the repurposed drug is deployed in combination with another antimalarial with similar PK properties, such as the elimination half-life, to avoid resistance selection. These considerations must be balanced against the critical need for an alternative to SP-AQ for chemoprevention interventions.

Beyond simply confirming that intervention coverage is critical for SMC’s public health impact, our results indicate that optimising drug properties likely to affect SMC’s coverage will be more important than optimising duration to improve population impact beyond what is currently observed with SP-AQ. Drug dosing frequency and concentration is typically chosen to maximise a drug’s effect whilst maintaining an acceptable safety profile, and allowing for sufficient adherence. For SMC candidate selection, however, we may consider a product with a reducedduration of protectioninfavour of increasedadherence. Safety on repeateddosing is of particular concern for drug use in repeated chemoprevention cycles, potentially requiring lower dosage than for other use cases. Furthermore, parental reluctance to allow a child to receive any drug has been identified as a significant hurdle to adherence with SP-AQ,^35^ and SP-AQ’s three-dose schedule and known side effects of nausea and vomiting are likely to be significant drivers of non-adherence. Greater effectiveness could be achieved by providing an intervention with the least frequent dosing regimen possible to achieve a minimum acceptable effect. For SMC and for other chemoprevention programs, such as perennial malaria chemoprevention, it will be critical to pursue child-friendly formulations as early as possible, as has been done for SP. Close collaboration between dr ug developers and regulatorybodies will be crucial for integrating early feedback on the appropriate balance between drug efficacy and campaign coverage.

Development pathways for new chemoprevention drugs may also need to be designed to engage communities early and frequently, to secure buy in, and to address barriers to coverage early on. Questions regarding optimal dosing and cycle frequency should be explored prior to phase two trials in collaboration with malaria control programs, to ensure that the drug regimens evaluated can then be feasibly deployed within a malaria control program. Patient reported outcome measures have also become widely accepted measures of treatment benefit and risk in therapeutic areas such oncology and ophthalmology, where medical treatment focuses on extending or improving quality of life.^36^ The inclusion of patient reported outcome measures in phase two and three chemoprevention trials could allow for community preferences for drug deployment to be identified prior to pilot studies, enabling selectionof dosingregimens most likely to maximise drug adherence and achieve adequate SMC coverage.

As in any modelling study, this study has its limitations. First, our three drug models may not capture the as-yet-unknown dynamics of new chemoprevention drugs. In particular, both dominant blood and liver stage models assume complete liver and blood stage parasite clearance, respectively, at the time of administration. This assumption may underplay the importance of a drug’s killing effect and should be reconsidered as new clinical evidence becomes available.

Second, our three drug models described varying mechanisms of action for one drug with a minimal tail effect, which may not accurately represent drug combination candidates. In particular, we recognise that our one-compartment PK/PD models cannot fully represent drug-drug interactions. And, critically, they are unable to capture the importance of combining drug candidates with matching half-lives to prevent parasite resistance. However, representingthe combinedaction of multiple drugs with a single model offers a balance between model complexity and the substantial unknowns regarding chemoprevention drug PK/PD properties. In the future, as more clinical evidence for chemoprevention candidates becomes available, multiple models may be necessary to capture the effects of drug synergisms and interactions.

Last, while we included PK/PD models within our larger population model, our estimates for next-generation SMC’s effectiveness were based on a single, individual-based model of malaria transmission dynamics. We also focused on predicting the isolated impact of an SMC candidate’s public health impact for a limited range of deployment modalities and malaria-endemic settings. As such, this study does not provide precise predictions of SMC’s likely impact within a malaria control program, but rather aims to identify guiding principles to inform clinical development and support candidate selection. Moreover, decisions made based on evidence from modelling should be accompanied with a solid understanding of intervention dynamics from early clinical studies and clinical trial data, as well as evidence of feasible levels of coverage and adherence from implementation studies, as discussed above.

In conclusion, this study provides evidence-based support for chemoprevention drug candidate design and selection for effectiveness of SMC programs. Results demonstrate the importance of selecting and evaluating chemoprevention drugs for their duration of protection first and treatment efficacy second. Results also highlight the need to match SMC cycle timing to the drug’s duration of protection, which would necessitate additional studies to assess the impact on acceptability, feasibility, and SMC coverage. Providing each eligible child with at least one chemoprevention cycle remains key, and drug candidates must be designed or selected for ease of administration, adherence, and community acceptance.

## Contributors

MAP conceived the study. LBM developed and performed analyses. LBM, JM, TM, NN, SK, and MAP validated the model, analyses, and results. LBM prepared figures and drafted the manuscript. All authors contributed to interpreting the results and making edits to the draft and final manuscript, and gave their final approval for publication.

## Data sharing

All code and datasets are available on GitHub (https://github.com/lydiab-mayer/modelling-smc-drug-properties/tree/v1.0.0).

## Data Availability

All code and datasets are available on GitHub

https://github.com/lydiab-mayer/modelling-smc-drug-properties/tree/v1.0.0

## Acknowledgements

This study was funded by the Bill & Melinda Gates Foundation (INV-002562 to MAP). TM and MAP acknowledge support from the Swiss National Science Foundation (SNF Professorship PP00P3_170702 and PP00P3_203450 to MAP). AMT is an employee of Medicines for Malaria Venture (MMV). We sincerely acknowledge all attendees of the 2021 convening ‘Malaria Prevention: Shaping Next-Gen Medical Interventions’ for their discussion and feedback. In addition, we wish to thank Jean-Luc Bodmer (Bill & Melinda Gates Foundation) and Scott Miller (previously Bill & Melinda Gates Foundation, now at Gates Medical Research Institute) for their contributions to this project and acknowledge support and advice from all members of the Disease Modelling Research unit of the Swiss Tropical and Public Health Institute, and Dr Lydia Burgert. We would like to thank Nadja Cereghetti for project management support. Calculations were performed at sciCORE (http://scicore.unibas.ch/) scientific computing centre at University of Basel.

## Appendix

### 1. Methods

#### 1.1. Mathematical transmission model

The potential public health impact of next-generation seasonal malaria chemoprevention (SMC) was predicted using an established and validated individual-based malaria transmission model, OpenMalaria (https://github.com/SwissTPH/openmalaria/wiki).^1^ OpenMalaria has multiple model components, each of which captures a different set of malaria epidemiologyand transmissionassumptions and has been fully described previously.^2–4^ Here we present an overview of the relevant dynamics for the model variant used in our study, which included an explicit mechanistic model of within-host parasite dynamics^5^ and incorporated flexible pharmacokinetic/pharmacodynamic (PK/PD) model for drug action.^2^

OpenMalaria generates discrete, stochastic simulations of malaria infection and transmission from individual human hosts, tracked in five-day time steps. This model, which was fully described again in Reiker and colleagues,^3^ with key components listed in table A1.1, and features: seasonally forced sub-models representing malaria parasite transmission within mosquitos; stochastic predictions of parasite densities within humans, determined by a malaria infection model and acquired immunity to asexual blood stage parasitaemia, and; the ability to track and distinguish between uncomplicated and severe episodes of clinical malaria, as well as direct and indirect malaria mortality. Together, these features enable detailed simulation of malaria public health outcomes for different interventions, such as the impact of malaria case management, drug-based chemoprevention interventions, and vector control.^6^

The within-host model variant adds a mechanistic feature of parasite dynamics within the human host, where each infection is modelled with a mechanistic *Plasmodium falciparum* parasite model adapted from Molineaux and colleagues,^5^ and incorporates full PK/PD models of intervention dynamics. This model mechanistically describes the time-course of asexual *Plasmodium falciparum* parasitaemia following a single inoculation within an individual human host. This time-course is driven by a parasite growth rate and by the effect of three different immune responses to parasitaemia within the human host – innate and cross-variant immunity, acquired and variant specific immunity, and acquired and cross-variant immunity – each of which acts to reduce the parasite growth rate. By using a malaria transmission model with this realistic mathematical, within-host model of asexual parasitaemia, we capture a medical intervention’s impact on both the time-course of asexual *P. falciparum* parasitaemia and changes to acquired immunity by modelling the drug’s impact on parasite growth.

**Table A1.1:**
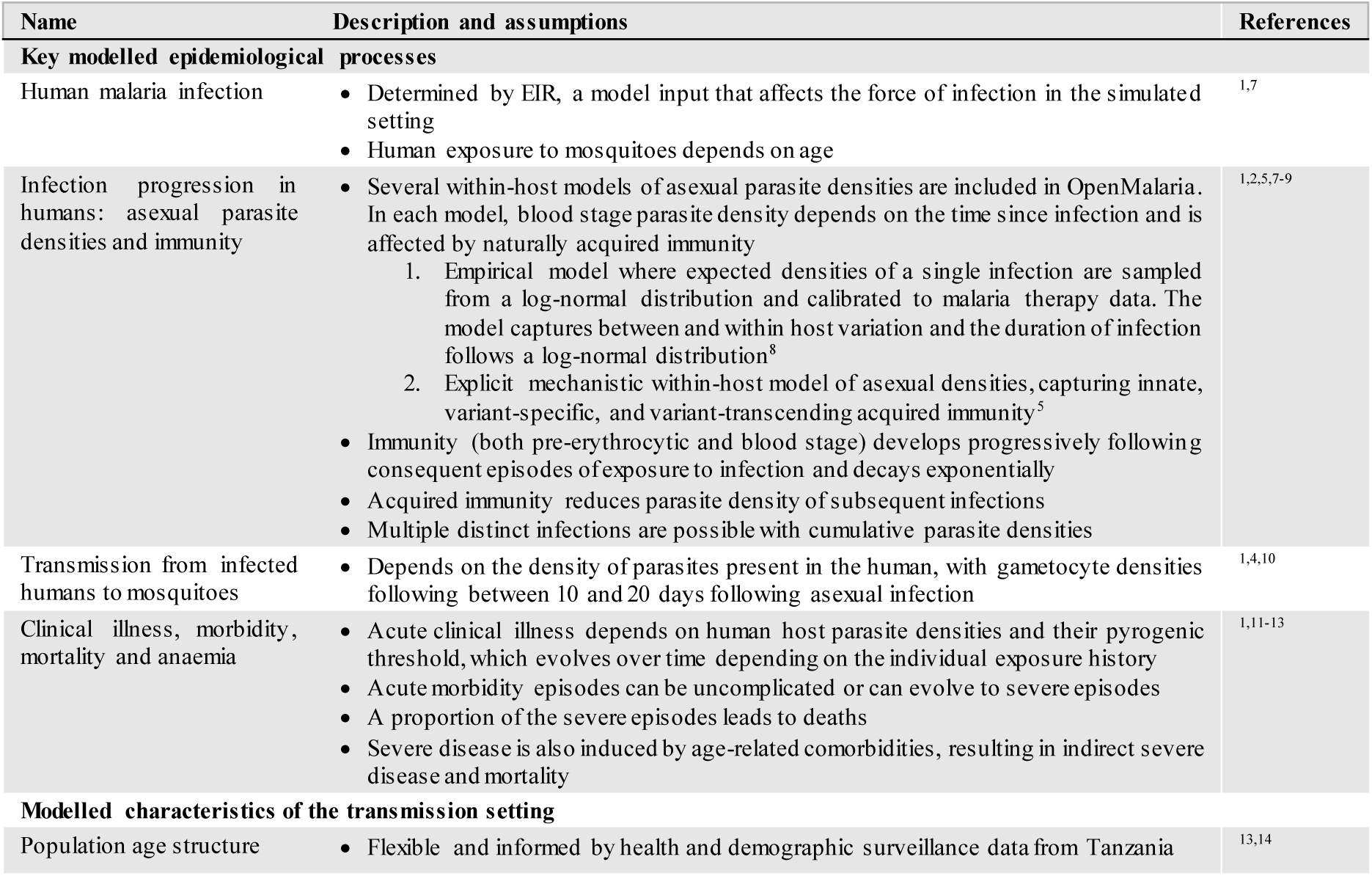
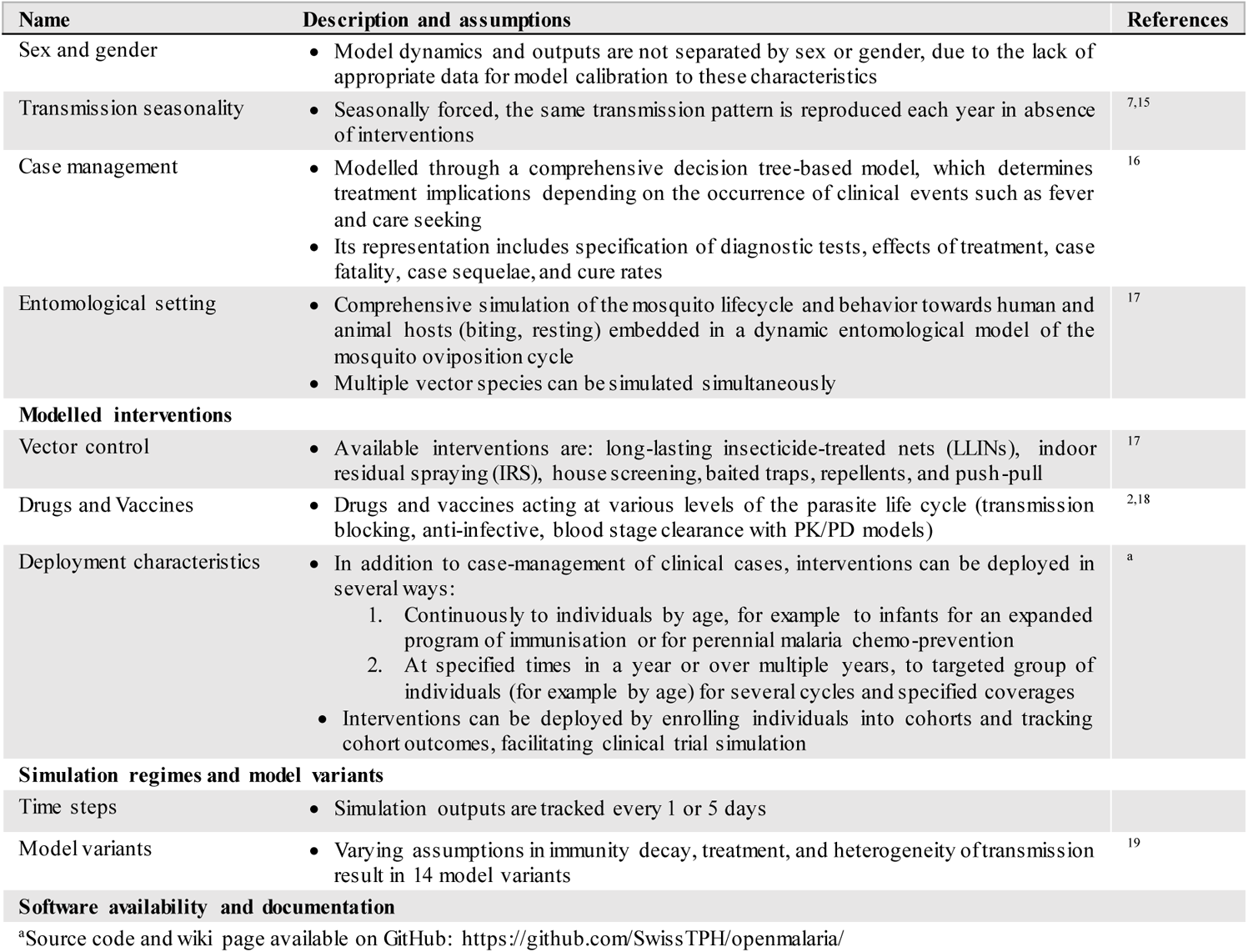
Summary of disease model characteristics, adapted from Golumbeanu et al. ^20^.

| Name | Description and assumptions | References |
| --- | --- | --- |
| **Key modelled epidemiological processes** |  |  |
| Human malaria infection | Determined by EIR, a model input that affects the force of infection in the simulated setting<br>Human exposure to mosquitoes depends on age | 1,7 |
| Infection progression in humans: asexual parasite densities and immunity | Several within-host models of asexual parasite densities are included in OpenMalaria. In each model, blood stage parasite density depends on the time since infection and is affected by naturally acquired immunity<br>Empirical model where expected densities of a single infection are sampled from a log-normal distribution and calibrated to malaria therapy data. The model captures between and within host variation and the duration of infection follows a log-normal distribution<sup>8</sup><br>Explicit mechanistic within-host model of asexual densities, capturing innate, variant-specific, and variant-transcending acquired immunity<sup>5</sup><br>Immunity (both pre-erythrocytic and blood stage) develops progressively following consequent episodes of exposure to infection and decays exponentially<br>Acquired immunity reduces parasite density of subsequent infections<br>Multiple distinct infections are possible with cumulative parasite densities | 1,2,5,7-9 |
| Transmission from infected humans to mosquitoes | Depends on the density of parasites present in the human, with gametocyte densities following between 10 and 20 days following asexual infection | 1,4,10 |
| Clinical illness, morbidity, mortality and anaemia | Acute clinical illness depends on human host parasite densities and their pyrogenic threshold, which evolves over time depending on the individual exposure history<br>Acute morbidity episodes can be uncomplicated or can evolve to severe episodes<br>A proportion of the severe episodes leads to deaths<br>Severe disease is also induced by age-related comorbidities, resulting in indirect severe disease and mortality | 1,11-13 |
| **Modelled characteristics of the transmission setting** |  |  |
| Population age structure | Flexible and informed by health and demographic surveillance data from Tanzania | 13,14 |
| Sex and gender | Model dynamics and outputs are not separated by sex or gender, due to the lack of appropriate data for model calibration to these characteristics |  |
| Transmission seasonality | Seasonally forced, the same transmission pattern is reproduced each year in absence of interventions | 7,15 |
| Case management | Modelled through a comprehensive decision tree-based model, which determines treatment implications depending on the occurrence of clinical events such as fever and care seeking<br>Its representation includes specification of diagnostic tests, effects of treatment, case fatality, case sequelae, and cure rates | 16 |
| Entomological setting | Comprehensive simulation of the mosquito lifecycle and behavior towards human and animal hosts (biting, resting) embedded in a dynamic entomological model of the mosquito oviposition cycle<br>Multiple vector species can be simulated simultaneously | 17 |
| **Modelled interventions** |  |  |
| Vector control | Available interventions are: long-lasting insecticide-treated nets (LLINs), indoor residual spraying (IRS), house screening, baited traps, repellents, and push-pull | 17 |
| Drugs and Vaccines | Drugs and vaccines acting at various levels of the parasite life cycle (transmission blocking, anti-infective, blood stage clearance with PK/PD models) | 2,18 |
| Deployment characteristics | In addition to case-management of clinical cases, interventions can be deployed in several ways:<br>Continuously to individuals by age, for example to infants for an expanded program of immunisation or for perennial malaria chemo-prevention<br>At specified times in a year or over multiple years, to targeted group of individuals (for example by age) for several cycles and specified coverages<br>Interventions can be deployed by enrolling individuals into cohorts and tracking cohort outcomes, facilitating clinical trial simulation | <sup>a</sup> |
| **Simulation regimes and model variants** |  |  |
| Time steps | Simulation outputs are tracked every 1 or 5 days |  |
| Model variants | Varying assumptions in immunity decay, treatment, and heterogeneity of transmission result in 14 model variants | 19 |
| **Software availability and documentation** |  |  |

<sup>a</sup>Source code and wiki page available on GitHub: https://github.com/SwissTPH/openmalaria/

#### 1.2. Scenarios and outcome measures

**Table A1.2:** Summary of simulation scenarios and outcome measures.

| Term | Scenario |
| --- | --- |
| **Case management** | Results for two levels of health system access for first-line treatment of malaria were captured:<br>A low probability of seeking care for clinical illness (10% within 14 days).<br>A high probability of seeking care for clinical illness (50% within 14 days).<br>Treatment was assumed to clear all blood stage infections for one time-step. |
| **Primary outcomes** | We measured next-generation SMC's impact on three relative outcomes in the target population:<br>**Clinical incidence reduction:** The percentage reduction in clinical incidence compared with a no intervention counterfactual, where incidence was defined as the number of new, uncomplicated malaria cases across the three-, four-, or five-month intervention period in the fifth year after deployment. Uncomplicated malaria was defined as an episode of symptomatic malaria, detectable by rapid diagnostic test with 94.2% specificity and a detection limit of 50 parasites per microlitre, where symptoms did not qualify as severe malaria.<br>**Prevalence reduction:** The percentage reduction in prevalence compared with a no intervention counterfactual, where prevalence was defined as the proportion of all malaria infections (detectable by rapid diagnostic test or not) at the end of the three-, four-, or five-month intervention period in the fifth year after deployment.<br>**Severe disease reduction:** The percent reduction in the number of severe malaria cases compared with a no intervention counterfactual, where severe cases of malaria were evaluated across the three-, four-, or five-month intervention period in the fifth year after deployment. An episode of severe malaria was an episode of symptomatic malaria, detectable by rapid diagnostic test with 94.2% specificity and a detection limit of 50 parasites per microlitre, with symptoms qualifying as severe malaria or with co-morbidities<sup>1</sup>.<br>All outcomes were evaluated in the intervention's target population: children aged three to 59 or three to 119 months. |
| **Seasonality** | Differences in SMC's performance were captured across types of seasonal transmission by reporting results for archetypal seasonal profiles, where approximately 70% of cases occurred within three or five months in a given year (figure A1.2). |
| **SMC cycles** | The importance of covering the entire length of a region's highest risk period to malaria infection was evaluated by varying the number of cycles of SMC: |

**Table A1.2: Summary of simulation scenarios and outcome measures**
|  |  |
| --- | --- |
| **Target population** | SMC was evaluated for two target populations:<br>• Children three to 59 months (5 years) old.<br>• Children three to 119 months (10 years) old. |
| **Transmission intensity** | SMC's performance was explored across a variety of malaria-endemic regions by providing results across a range of transmission intensities, from low (8%) to high (39%) annual baseline annual prevalence rate of *P. falciparum* patent infections (detectable by rapid diagnostic test with 94.2% specificity and a detection limit of 50 parasites per microlitre) in children aged two to ten years of age ($PfPR_{2-10}$) compared with the no SMC intervention counterfactual. |

**Figure A1.2:**
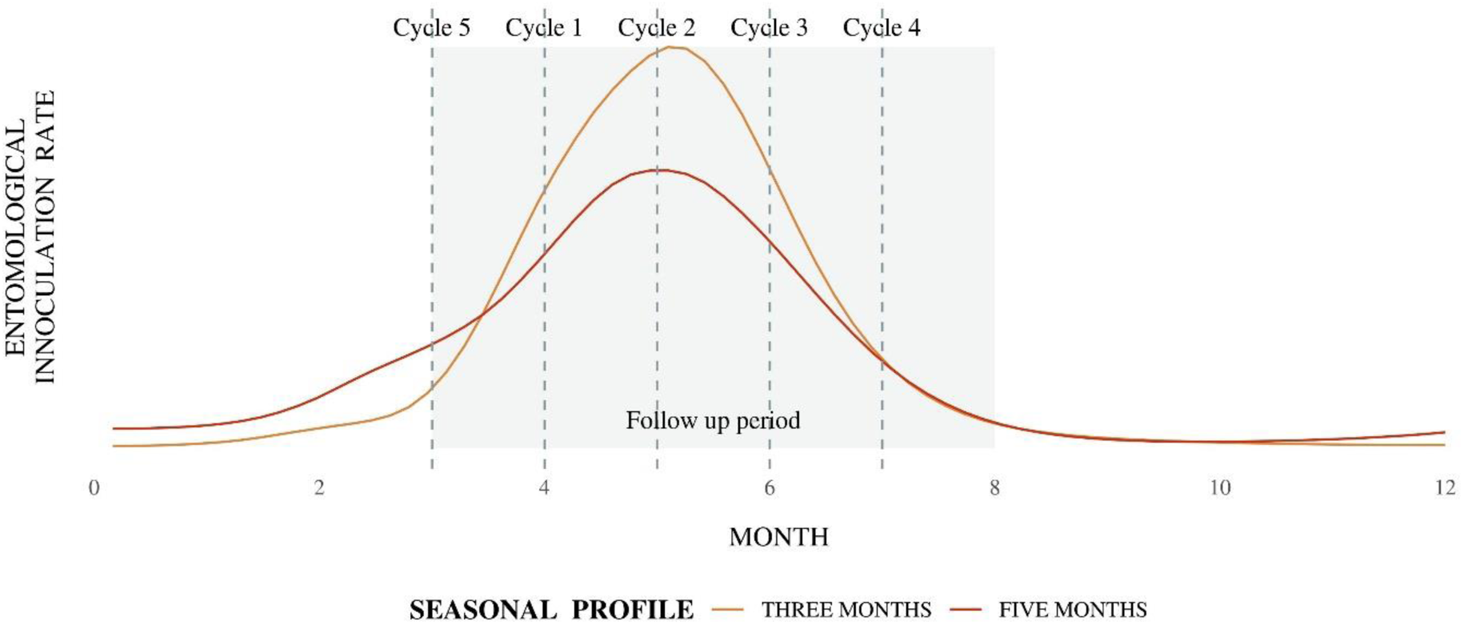
Seasonality profiles and SMC cycle timing.

#### 1.3. Intervention dynamics

##### Next-generation SMC with blood stage activity only

To model next-generation SMC drugs with blood stage activity only, we used a one-compartment pharmacokinetic/pharmacodynamic (PK/PD) model to describe a next-generation SMC drug’s killing effect at time *t* as

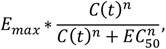

where *C(t)* is the drug’s concentration in mg/liter, *E_max_* the maximum parasite killing rate per day, *EC_50_* the concentration at which 50% of the maximum killing rate occurs in mg/liter, and *n* the slope of the drug’s dose response curve. ^21^ The drug’s concentration decays over time *t* with

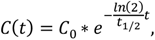

Where *C_0_* is the drug’s concentration at time zero and *t_1/2_* is the elimination half-life.

For simplicity, given that pharmacokinetic properties for next-generation SMC are still unknown, we assumed a piperaquine-like treatment schedule with a single dose administered daily for three days at a weight-based dosing schedule of 18 mg/kg.^22^ Parameter values for the model’s volume of distribution and *EC_50_* were selected to represent piperaquine-like behavior,^23^ as detailed in table 1.

We captured uncertainty around next-generation SMC’s pharmacological properties by varying key parameters for drug efficacy and duration. To capture uncertainty about drug efficacy, we varied the *E_max_* between two and 30. To represent uncertainty in a drug’s duration of protection, we ranged the elimination half-life *t_1/2_* between five and 40 days. In this one-compartment PK/PD, uncertainty in drug duration could equally have been represented by ranging the EC_50_. We varied the EC_50_ in our initial analyses and elected to present results for a fixed value. This modellingdecisionwas made in order to limit the complexityof performingoptimisationontwo parameters for duration. The slope *n* was also varied in initial analyses but was found to have minimal contribution to model outcomes, which were presented for a slope of six.

The PK component of this model has only one compartment and assumes instantaneous absorption and, as such, may not reflect the complexityof a chemopreventiondrug’s exposure-response relationship. However, this simple approach to modelling SMC captured a next-generation drug’s potential for activity against blood stage parasites without making complex assumptions about that drug’s as-yet-unknown PK properties.

##### Next-generation SMC with dominant blood stage activity and initial, complete liver stage clearance

Some next-generation SMC candidates are likely to be partially active against liver stage parasites. For this reason, our ‘dominant blood stage activity’ model deployed the PK/PD model described above with the addition of one time-step of liver stage parasite clearance from the start of SMC administration.

##### Next-generation SMC with dominant liver stage activity and initial, complete blood stage clearance

To capture next-generation SMC’s likely protective effect against liver stage parasites, we modelled the intervention’s protective effect as a probability of preventing liver stage infection that decays over time, in combination with five days of blood and liver stage parasite clearance from the start of SMC administration. This approach, which was based on a previously calibrated model of SP-AQ,^24^ has two key parameters: duration of protection, driven by the drug’s elimination half-life and *EC_50_*, and; initial probability of preventing infection, driven by the drug’s maximal effect *E_max_*. By modelling the intervention’s protective effect over time, we could translate a drug’s pharmacokinetics or pharmacodynamics to its expected public health impact. As for next-generation SMC with blood stage activity, we modelled a range of values for these key characteristics (table 1).

##### In-silico clinical trial modelling for the standard-of-care

We assessed each model’s ability to accurately represent the dynamics of SP-AQ through in-silico clinical trial modelling, where we replicated SP-AQ’s protective efficacy against clinical cases from a randomised non-inferiority trial of dihydroartemisinin-piperaquine (DHA-PPQ) to SP-AQ, conducted between 2009 and 2010 in Burkina Faso by Zongo and colleagues.^22^ Adapting Burgert and colleagues’ previous parameterization of SP-AQ to this same trial,^24^ using the following approach:

1. Protective efficacy data for SP-AQ against clinical cases was extracted from figure 3 of Zongo and colleagues, and the trial was re-built in OpenMalaria using the settings described in table A1.3.
2. For each of the three mechanism-of-action models, we used latin-hypercube sampling to generate 500 samples of parameter values within the ranges described in table 1.
3. For each potential mechanism-of-action and for each parameter sample, we ran ten stochastic replicates of the in-silico clinical trial with OpenMalaria, simulating both the intervention arm of the trial and a no-intervention control arm.
4. Protective efficacy was calculated per time-step as

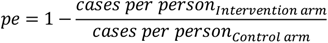

where clinical cases were detectable by rapid diagnostic test.
5. Protective efficacy for each potential mechanism-of-action was considered to be sufficiently close to that of SP-AQ if the residual sum of squares (RSS) was within 0·1 standard deviations of the minimum RSS.

**Table A1.3:** OpenMalaria settings to perform an in-silico clinical trial with SP-AQ.

| Parameter | Value | Reference |
| --- | --- | --- |
| Mosquito species | *Anopheles funestus*: 42% indoor biting and 31% outdoor biting<br>*Anopheles gambiae*: 14% indoor biting and 13% outdoor biting | 25 |
| Total entomological inoculation rate (EIR) | 350 for in-silico modelling of blood stage mechanism-of-actions<br>154 for in-silico modelling of liver stage mechanism-of-actions | .. |
| **Malaria seasonality (as monthly EIR)** | *Anopheles funestus*: 23 (Aug), 16 (Sep), 0 (Oct), 3.1 (Nov)<br>*Anopheles gambiae*: 40 (Aug), 50 (Sep), 16 (Oct), 6.4 (Nov) | 25 |
| **Access to treatment** | Effective coverage within 14-days of 50%, represented in OpenMalaria as 24.12% over 5-day time-steps | 26,27 |
| **Clinical malaria treatment pathway** | First-line treatment with artemeter-lumefantrine<br>Second-line treatment with quinine | 27 |
| **Insecticide treated net coverage** | 14% between 2006 and 2008, increased to 27% during the trial period | 22,27 |
| **SMC timing** | Cycles on 15-Aug, 15-Sep, and 15-Oct in 2009<br>Follow-up on 1-Sep, 1-Oct, and 1-Nov in 2009 | 22 |

#### 1.4. Simulation and statistical analysis

Following our previous methodology^20^ and as shown in figure A1.3, our predictive target product profile framework began with simulation from the individual-based malaria transmission model over a discrete sample of input intervention parameters. 1000 samples were uniformly generated with Latin hypercube sampling and, for each sample, simulations were generated with 5 stochastic replicates. Following simulation, we used the *hetgp* package^28^ to fit a heteroskedastic Gaussian Process regression model to emulate the relationship between input intervention parameters and their corresponding simulated outcomes. This step trained a computationally light model emulator of the individual-based malaria transmission model’s complex, deterministic dynamics. Emulator performance was assessed using the R-squared correlation coefficient to evaluate correlation between true and predicted outcomes against a 10% hold-out set.

We then undertook a nonparametric variance-based sensitivity analysis of our model results to identify the extent to which a small change in an SMC drug’s properties contributed to a change in its effectiveness. This was performed with the sensitivity package^29^, using the Sobol-Janssen method^30^ to compute Sobol total-order indices for two uniform samples of 50 000 input intervention parameters, generated with Latin hypercube sampling with 1000 bootstrap replicates. Sobol total-order indices represent the contribution of a given input variable to the variance of the output, called total-order or total-effect because this contribution includes any contribution to the variance by interactions with other input variables.

To identify desirable product characteristics for SMC, we linked a desired public health outcome with its required minimum intervention properties using an optimisation grid search. This was performed for blood stage models only by evaluating emulator predictions of public health outcomes on a sample of intervention properties (10 000 samples generated with Latin hypercube sampling) and identifying the estimated minimum property value whose 95% prediction interval was above the given target reduction in both clinical incidence and severe disease. This minimum was then aggregated by calculating the most conservative (maximum) value across outcomes (clinical incidence and severe disease reduction measured across the intervention period), SMC deployments (three, four, and five monthly cycles of SMC in a given year surrounding peak seasonality) and levels for other model parameters (elimination half-life and E_max_).All analyses were conducted in R.^31^

**Figure A1.3:**
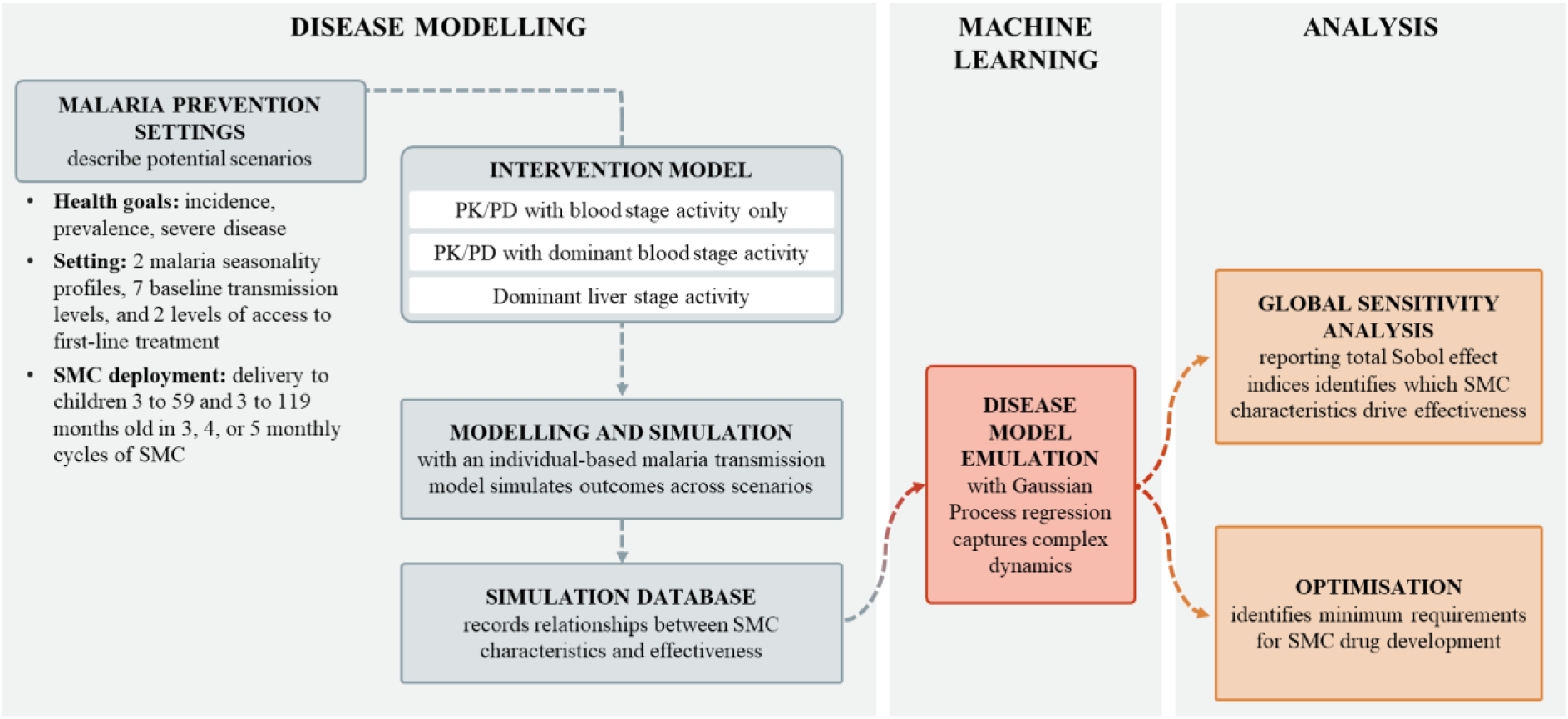
Evidence generation framework combining individual-based malaria transmission modelling with statistical methods to support the identification of minimum necessary SMC product characteristics.

### 2. Additional results

**Figure A2.1:**
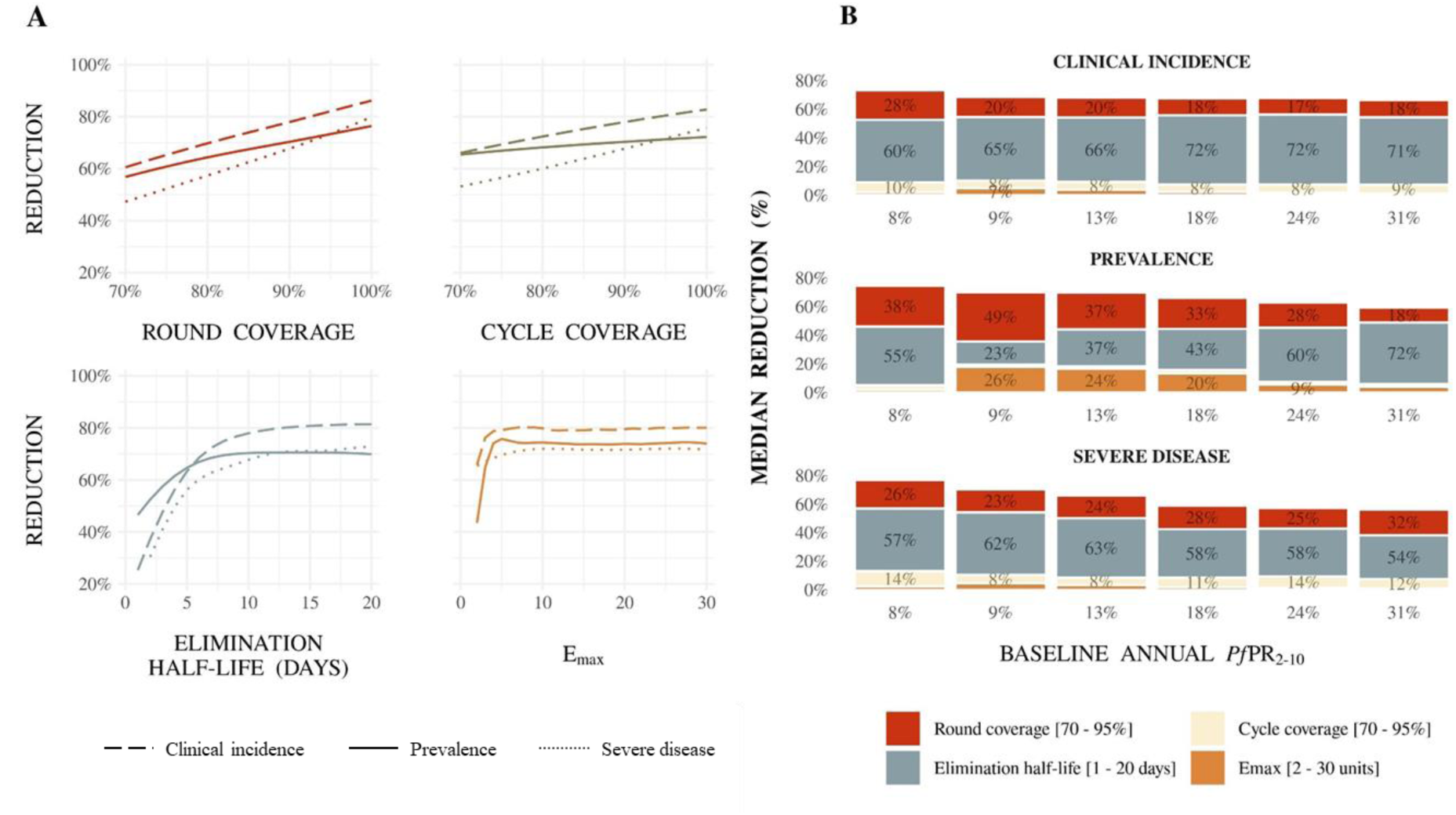
For SMC with blood stage activity only, (A) illustrative Gaussian Process regression emulator predictions for the relationship between SMC properties and clinical incidence, prevalence, and severe disease reduction, and (B) drivers of impact on predicted clinical incidence, prevalence, and severe disease reduction for SMC in children three to 59 months of age, compared with a no intervention counterfactual. (A) Results show 50% access to first-line treatment within 14 days and a five-month seasonal profile withbaseline annual *Pf*PR_2-10_ of 18% when SMC was deployed in four monthly cycles to children aged three to 59 months. Each panel shows the predicted reduction when all performance characteristics but the parameter of interest were held constant at: 90% round coverage, 90% cycle coverage, elimination half-life of 10 days, and a maximum parasite killing rate of 3.45 units. (B) Bars indicate the total Sobol effect indices for key model parameters. Indices can be interpreted as the proportion of variation in the outcome attributable to a given change in each variable, along with its interactions with other variables. Bar heights indicate the median expected reduction across the modelled parameter range. Results are shown across prevalence settings (horizontal axis, baseline annual *Pf*PR_2-10_) for an archetypal scenario of 50% access to first-line treatment within 14 days with a five-month seasonal profile, where SMC was deployed in four monthly intervals in a given year surrounding peak seasonality.

**Figure A2.2:**
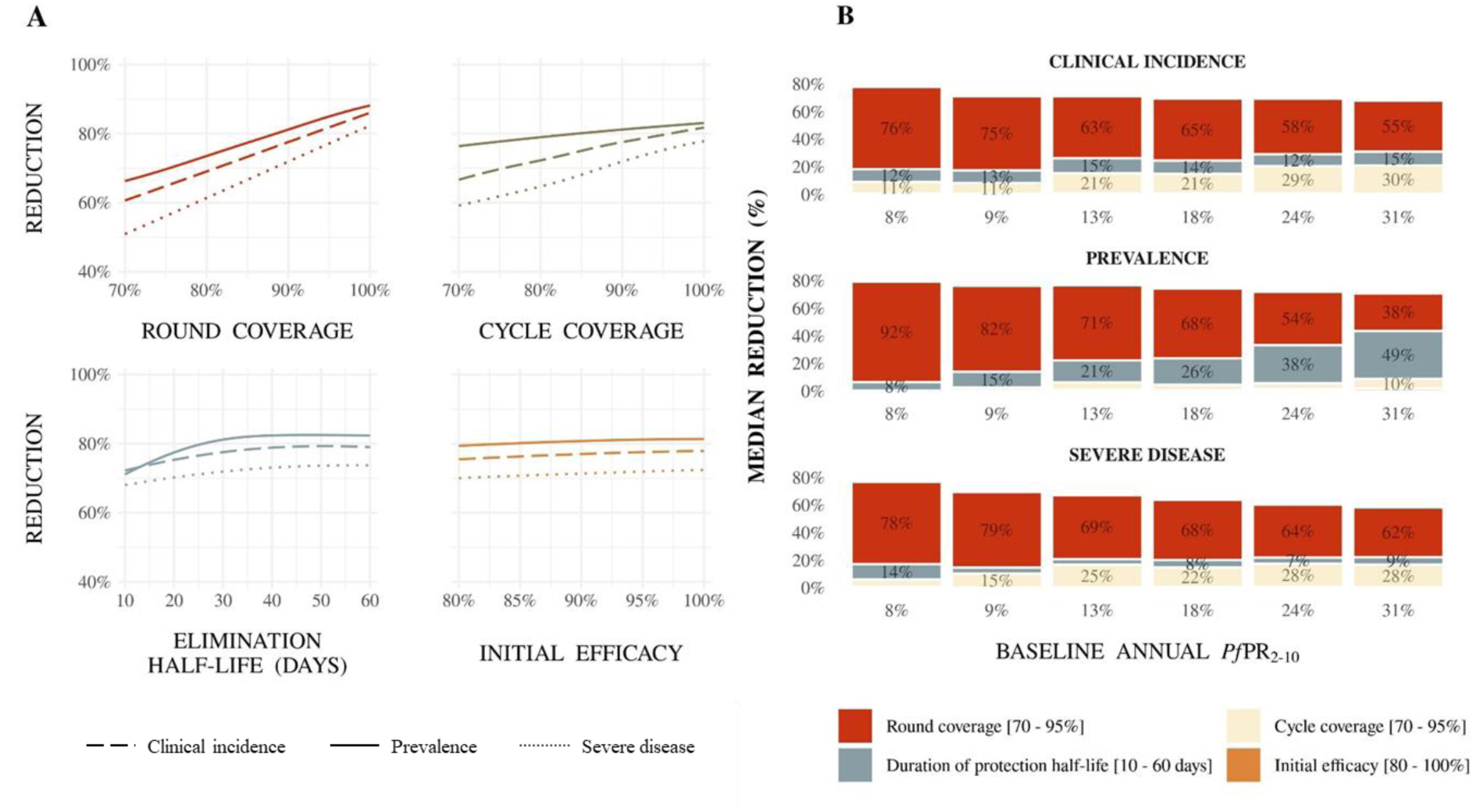
For SMC with dominant liver stage activity and initial, complete blood stage clearance, (A) illustrative Gaussian Process regression emulator predictions for the relationship between SMC properties and clinical incidence, prevalence, and severe disease reduction, and (B) drivers of impact on predicted clinical incidence, prevalence, and severe disease reduction for SMC in children three to 59 months of age, compared with a no intervention counterfactual. (A) Results show 50% access to first-line treatment within 14 days and a five-month seasonal profile with baseline annual *Pf*PR_2-10_ of 18% when SMC was deployed in four monthly cycles to children aged three to 59 months. Each panel shows the predicted reduction when all performance characteristics but the parameter of interest were held constant at: 90% round coverage, 90% cycle coverage, duration of protection half-life of 30 days, and initial efficacy of 95%. (B) Bars indicate the total Sobol effect indices for key model parameters. Indices can be interpreted as the proportion of variation in the outcome attributable to a given change in each variable, along with its interactions with other variables. Bar heights indicate the median expected reduction across the modelled parameter range. Results are shown across prevalence settings (horizontal axis, baseline annual *Pf*PR_2-10_) for an archetypal scenario of 50% access to first-line treatment within 14 days with a five-month seasonal profile, where SMC was deployed in four monthly intervals in a given year surrounding peak seasonality.

**Figure A2.3:**
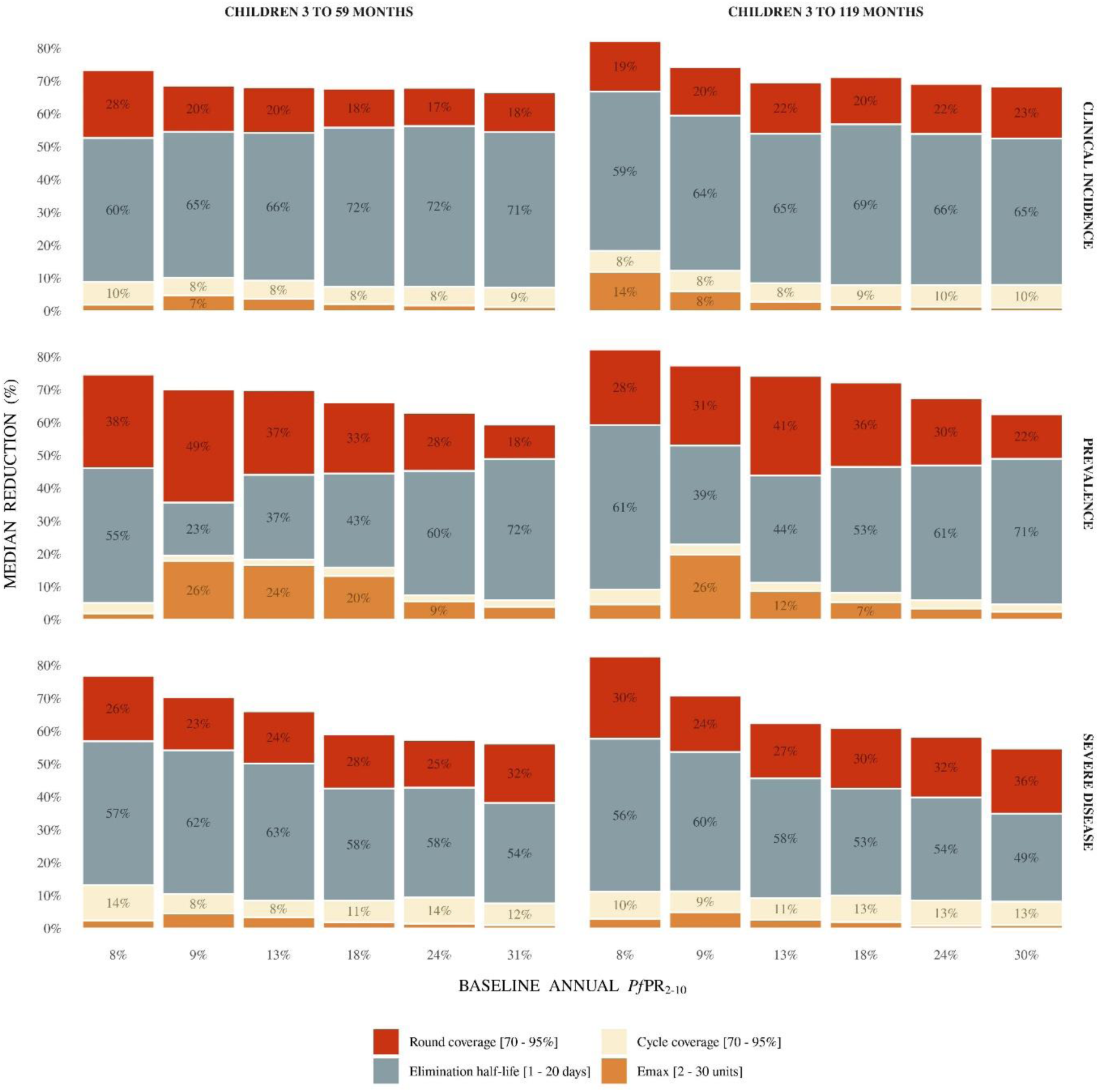
For SMC with blood stage activity only, drivers of impact on predicted levels of reduction of clinical incidence, prevalence, and severe disease in two scenarios – delivered to children aged three to 59 months or three to 119 months compared with the no intervention counterfactual. Bars indicate the total Sobol effect indices for key model parameters. Indices can be interpreted as the proportion of variation in the outcome attributable to a given change in each variable, along with its interactions with other variables. Bar heights indicate the median expected reduction across the modelled parameter range. Results are shown across prevalence settings (horizontal axis, annual baseline *Pf*PR_2-10_) for a scenario with 50% access to first-line treatment and a five-month seasonal profile where SMC was deployed in four monthly intervals in a given year.

**Figure A2.4:**
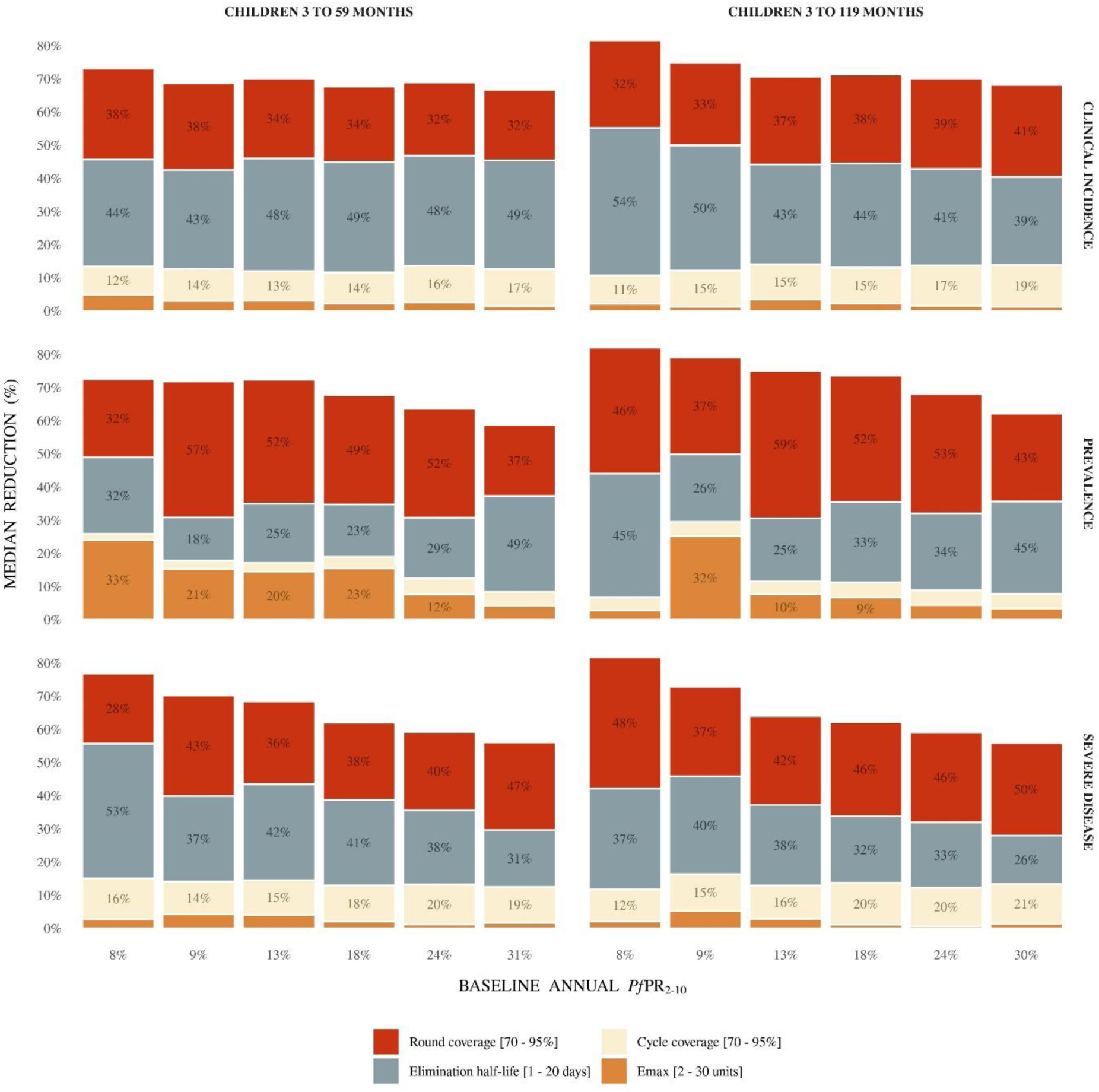
For SMC withdominant bloodstage activityandinitial, completeliver stage clearance, drivers of impact on predicted levels of reduction of clinical incidence, prevalence, and severe disease in two scenarios – delivered to children aged three to 59 months or three to 119 months compared with the no intervention counterfactual. Bars indicate the total Sobol effect indices for key model parameters. Indices can be interpreted as the proportion of variation in the outcome attributable to a given change in each variable, along with its interactions with other variables. Bar heights indicate the median expected reduction across the modelled parameter range. Results are shown across prevalence settings (horizontal axis, annual baseline *Pf*PR_2-10_) for a scenario with 50% access to first-line treatment and a five-month seasonal profile where SMC was deployed in four monthly intervals in a given year.

**Figure A2.5:**
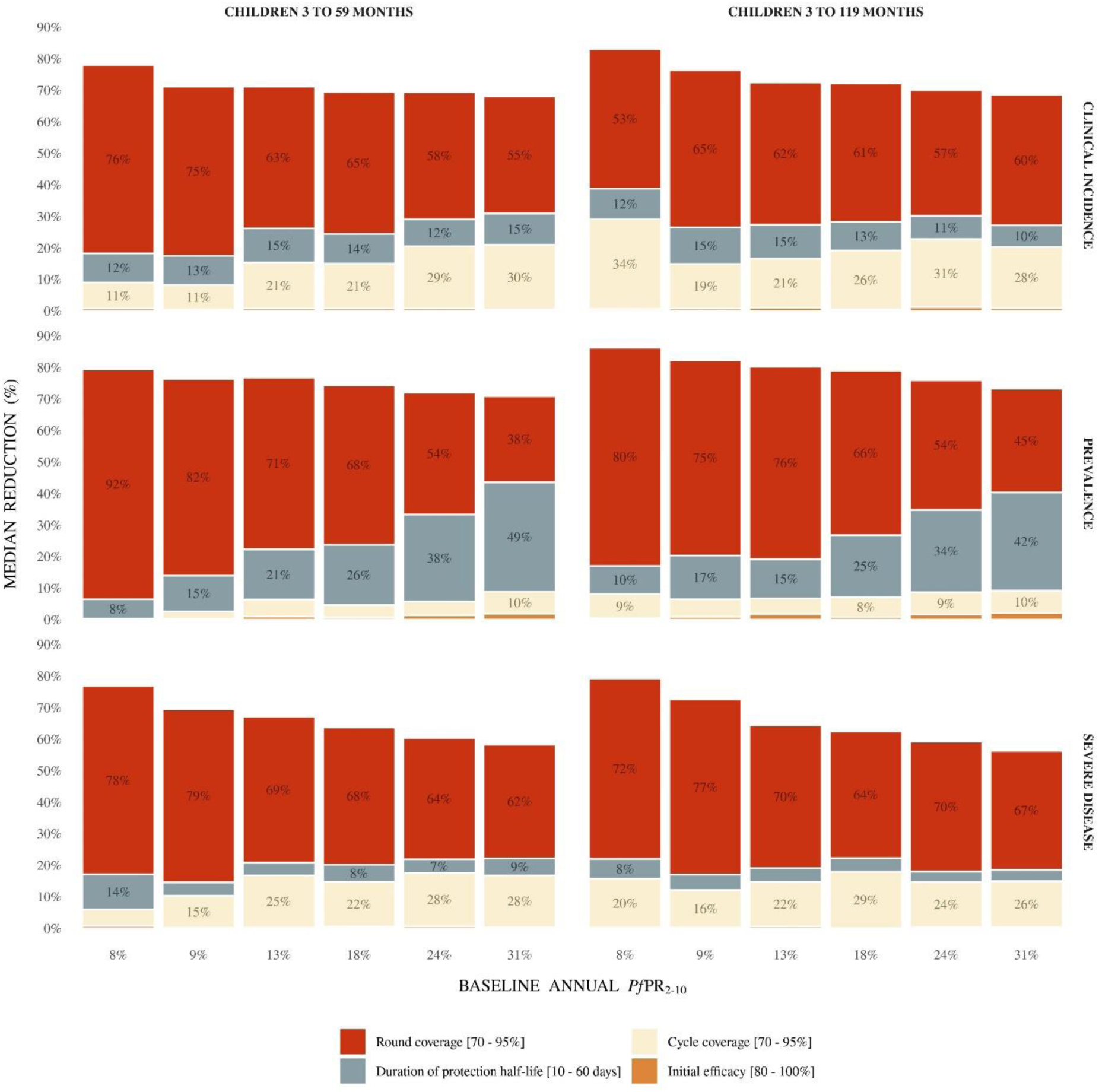
For SMC with dominant liver stage activity and initial, complete blood stage clearance, drivers of impact on predicted levels of reduction of clinical incidence, prevalence, and severe disease in two scenarios – delivered to children aged three to 59 months or three to 119 months compared with the no intervention counterfactual. Bars indicate the total Sobol effect indices for key model parameters. Indices can be interpreted as the proportion of variation in the outcome attributable to a given change in each variable, along with its interactions with other variables. Bar heights indicate the median expected reduction across the modelled parameter range. Results are shown across prevalence settings (horizontal axis, annual baseline *Pf*PR_2-10_) for a scenario with 50% access to first-line treatment and a five-month seasonal profile where SMC was deployed in four monthly intervals in a given year.

**Figure A2.6:**
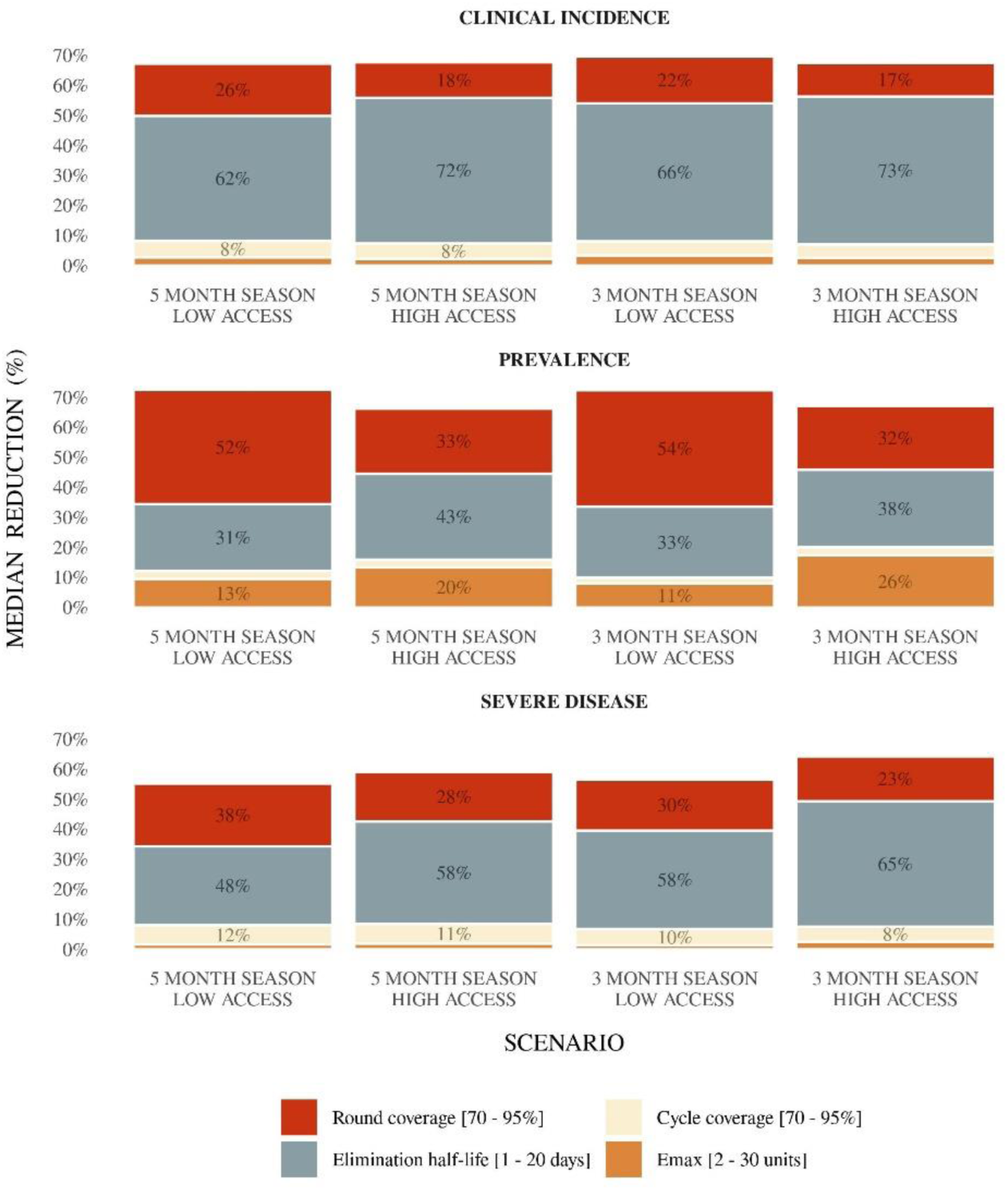
For next-generation SMC with blood stage activity only, drivers of impact on predicted levels of reductionofclinical incidence, prevalence, and severe disease for SMC representing four scenarios – low (10%) and high (50%) levels of access to malaria treatment, and three- and five-month seasonal profiles – compared with the no intervention counterfactual. Bars indicate the total Sobol effect indices for key model parameters. Indices can be interpreted as the proportion of variation in the outcome attributable to a given change in each variable, along with its interactions with other variables. Bar heights indicate the median expected reduction across the modelled parameter range. Results are shown for a scenario where SMC is deployed four times at monthly intervals in a given year in children aged three to 59 months, and where the annual baseline *Pf*PR_2-10_ varied with seasonal profile and access from 14% to 24%.

**Figure A2.7:**
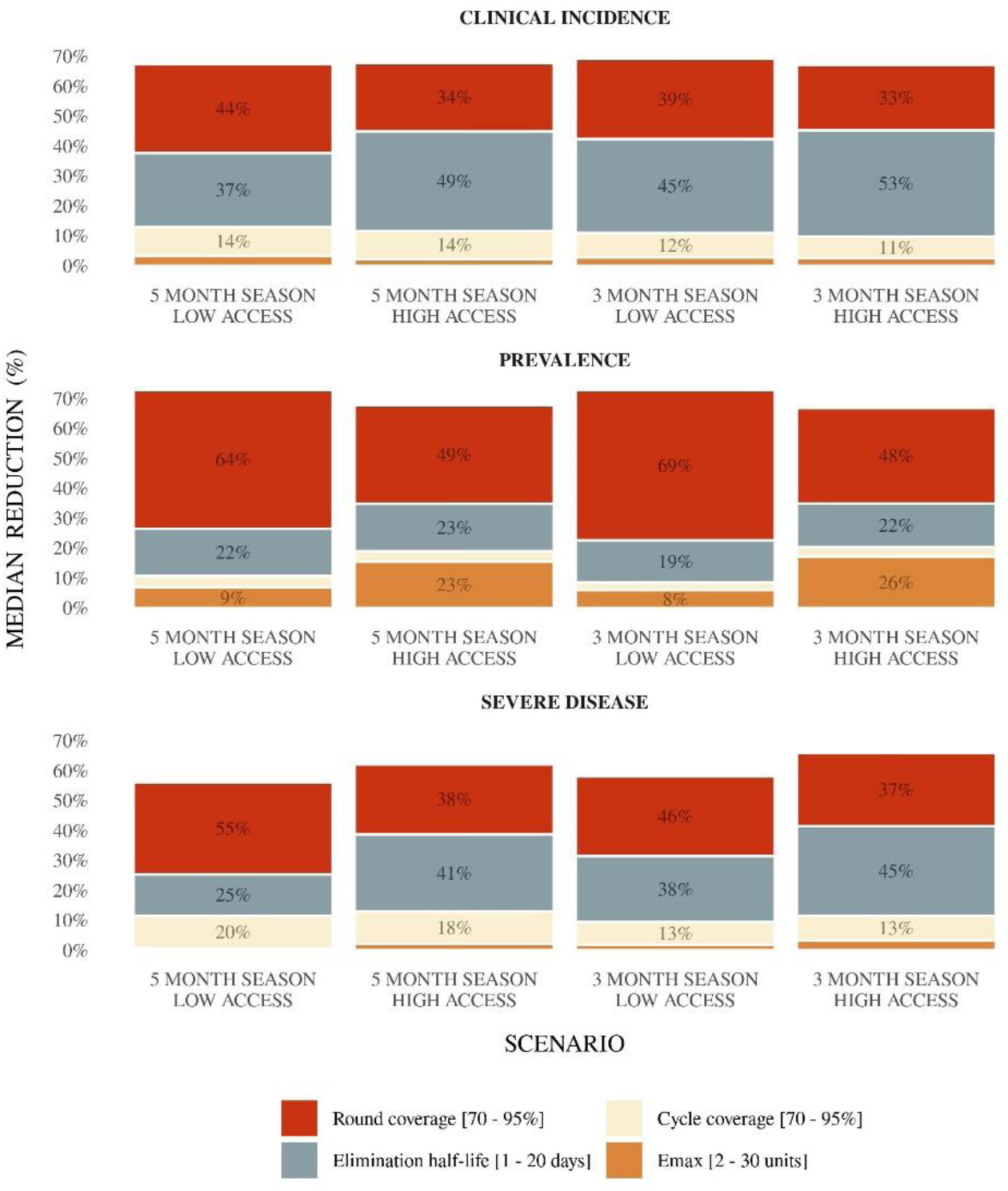
For next-generation SMC with dominant blood stage activity and initial, complete liver stage clearance, drivers of impact on predicted levels of reduction of clinical incidence, prevalence, and severe disease for SMC representing four scenarios – low (10%) and high (50%) levels of access to malaria treatment, and three- and five-month seasonal profiles –compared with the no intervention counterfactual. Bars indicate the total Sobol effect indices for key model parameters. Indices can be interpreted as the proportion of variation in the outcome attributable to a given change in each variable, along with its interactions with other variables. Bar heights indicate the median expected reduction across the modelled parameter range. Results are shown for a scenario where SMC is deployed four times at monthly intervals in a given year in children aged three to 59 months, and where the annual baseline *Pf*PR_2-10_ varied with seasonal profile and access from 14% to 24%.

**Figure A2.8:**
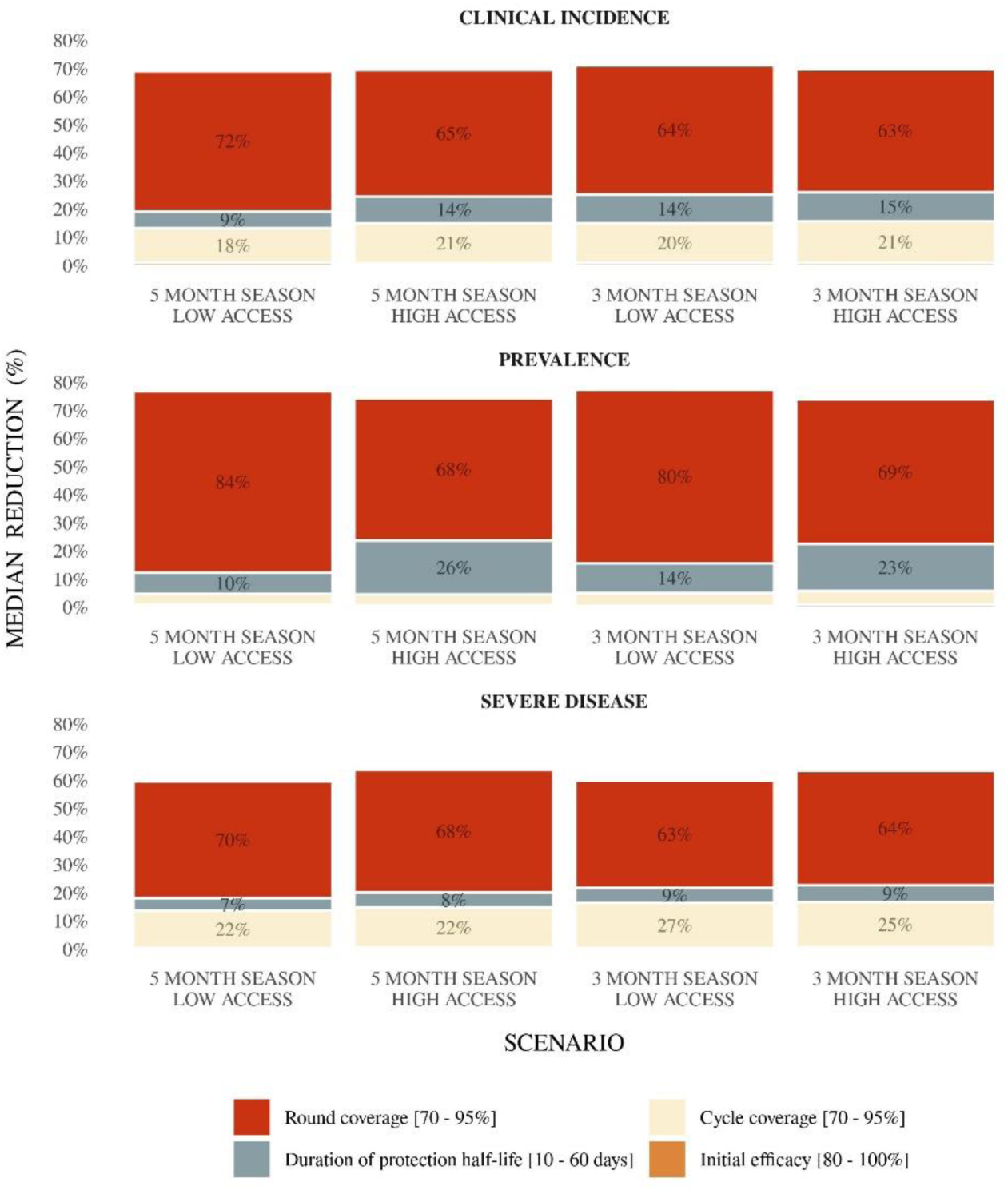
For next-generation SMC with dominant liver stage activity and initial, complete blood stage clearance, drivers of impact on predicted levels of reduction of clinical incidence, prevalence, and severe disease for SMC representing four scenarios – low (10%) and high (50%) levels of access to malaria treatment, and three- and five-month seasonal profiles –compared with the no intervention counterfactual. Bars indicate the total Sobol effect indices for key model parameters. Indices can be interpreted as the proportion of variation in the outcome attributable to a given change in each variable, along with its interactions with other variables. Bar heights indicate the median expected reduction across the modelled parameter range. Results are shown for a scenario where SMC is deployed four times at monthly intervals in a given year in children aged three to 59 months, and where the annual baseline *Pf*PR_2-10_ varied with seasonal profile and access from 14% to 24%.

**Figure A2.9:**
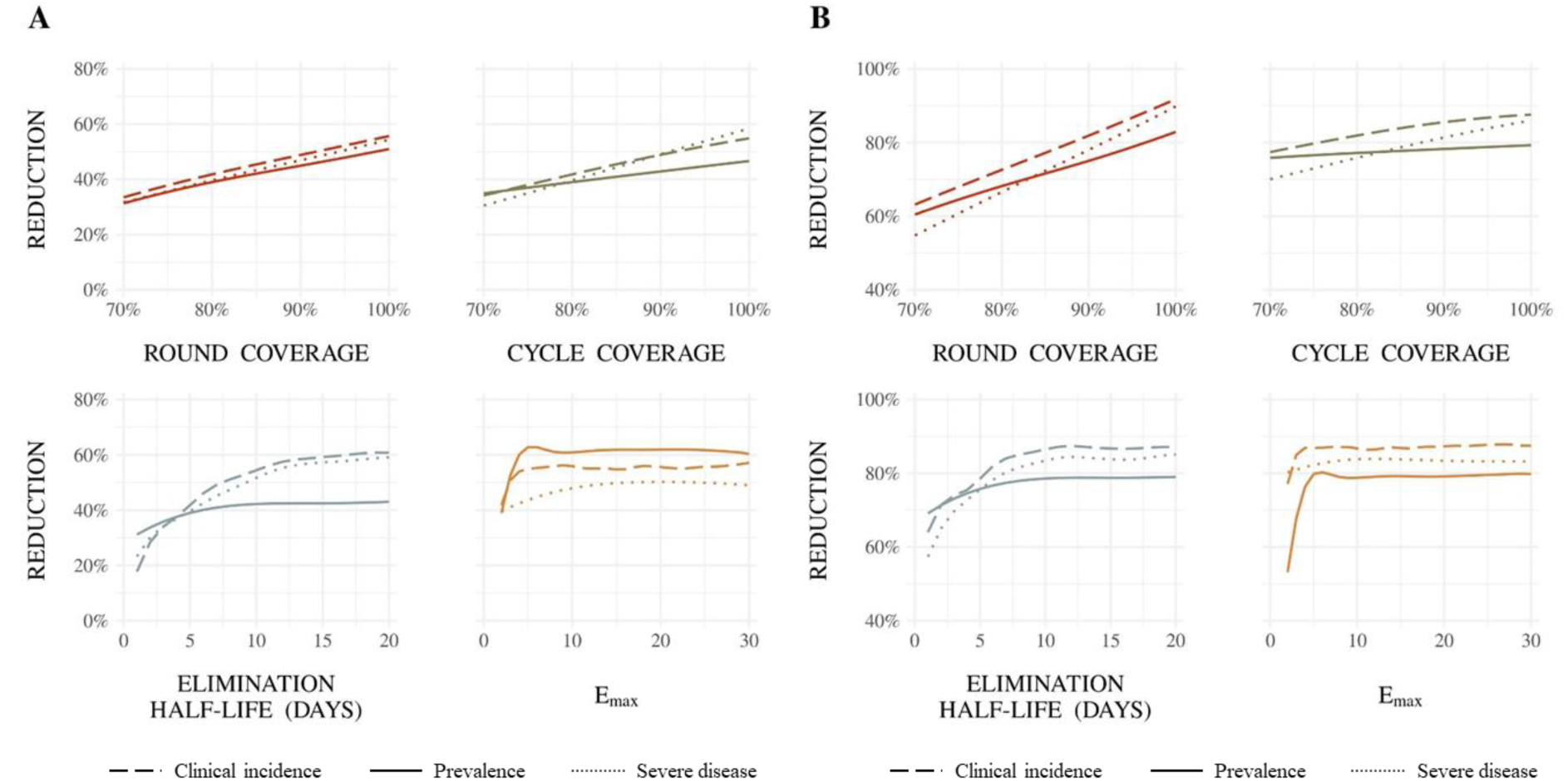
For SMC with dominant blood stage activity and initial, complete liver stage clearance, (A) illustrative Gaussian Process regression emulator predictions for the relationship between SMC properties and their direct effects on clinical incidence, prevalence, and severe disease reduction for a drug profile with low coverage and low protective efficacy, and (B) illustrative Gaussian Process regression emulator predictions for the relationship between SMC properties and their indirect effects on clinical incidence, prevalence, and severe disease reduction for a drug profile with high coverage and high protective efficacy. (A) Results show 50% access to first-line treatment within 14 days and a five-month seasonal profile with baseline annual *Pf*PR_2-10_ of 18% when SMC was deployed in four monthly cycles to children aged three to 59 months. Each panel shows the predicted reduction when all performance characteristics but the parameter of interest were held constant at: 80% round coverage, 80% cycle coverage, an elimination half-life of 5 days, and a maximum parasite killing rate of 2 units. (B) Results show 50% access to first-line treatment within 14 days and a five-month seasonal profile with baseline annual *Pf*PR_2-10_ of 18% when SMC was deployed in four monthly cycles to children aged three to 59 months. Each panel shows the predicted reduction – evaluated in SMC naïve children aged 60 to 119 months in the first intervention year – when all performance characteristics but the parameter of interest were held constant at: 95% round coverage, 95% cycle coverage, an elimination half-life of 15 days, and a maximum parasite killing rate of 10 units.

**Figure A2.10:**
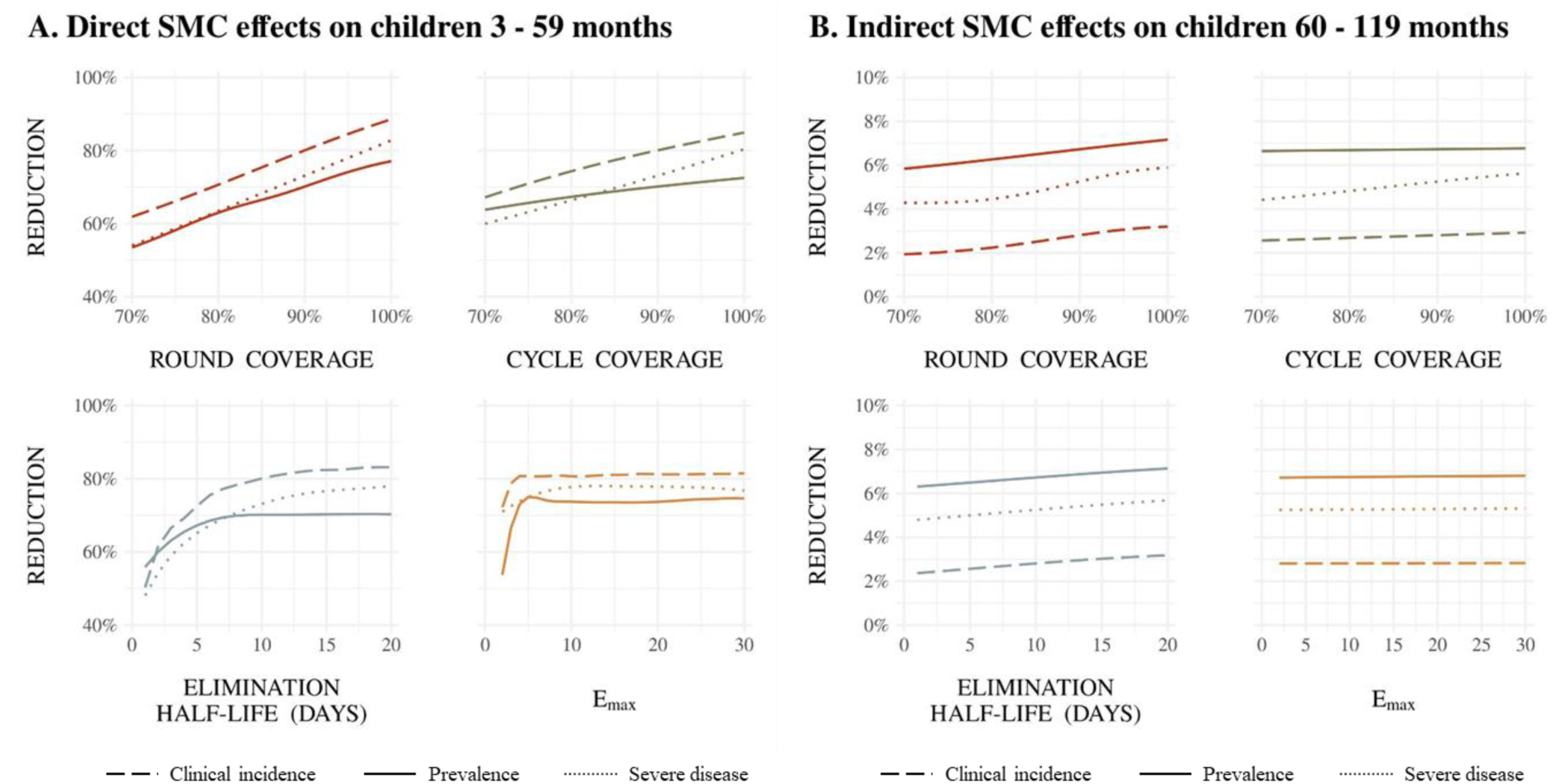
For SMC with dominant blood stage activity and initial, complete liver stage clearance, (A) illustrative Gaussian Process regression emulator predictions for the relationship between SMC properties and their direct effectsonclinical incidence, prevalence, andsevere diseasereductioninchildrenagedthree to 59 months receiving SMC, and (B) illustrative Gaussian Process regression emulator predictions for the relationship between SMC properties and their indirect effects on clinical incidence, prevalence, and severe disease reduction in children aged 60 to 119 months not receiving SMC. (A) Results show 50% access to first-line treatment within 14 days and a five-month seasonal profile with baseline annual *Pf*PR_2-10_ of 18% when SMC was deployed in four monthly cycles to children aged three to 59 months. Each panel shows the predicted reduction – evaluated in the first intervention year – when all performance characteristics but the parameter of interest were held constant at: 90% round coverage, 90% cycle coverage, an elimination half-life of 10 days, and a maximum parasite killing rate of 3.45 units. (B) Results show 50% access to first-line treatment within 14 days and a five-month seasonal profile with baseline annual *Pf*PR_2-10_ of 18% when SMC was deployed in four monthly cycles to children aged three to 59 months. Each panel shows the predicted reduction – evaluated in SMC naïve children aged 60 to 119 months in the first intervention year – when all performance characteristics but the parameter of interest were held constant at: 90% round coverage, 90% cycle coverage, an elimination half-life of 10 days, and a maximum parasite killing rate of 3.45 units.

**Figure A2.11:**
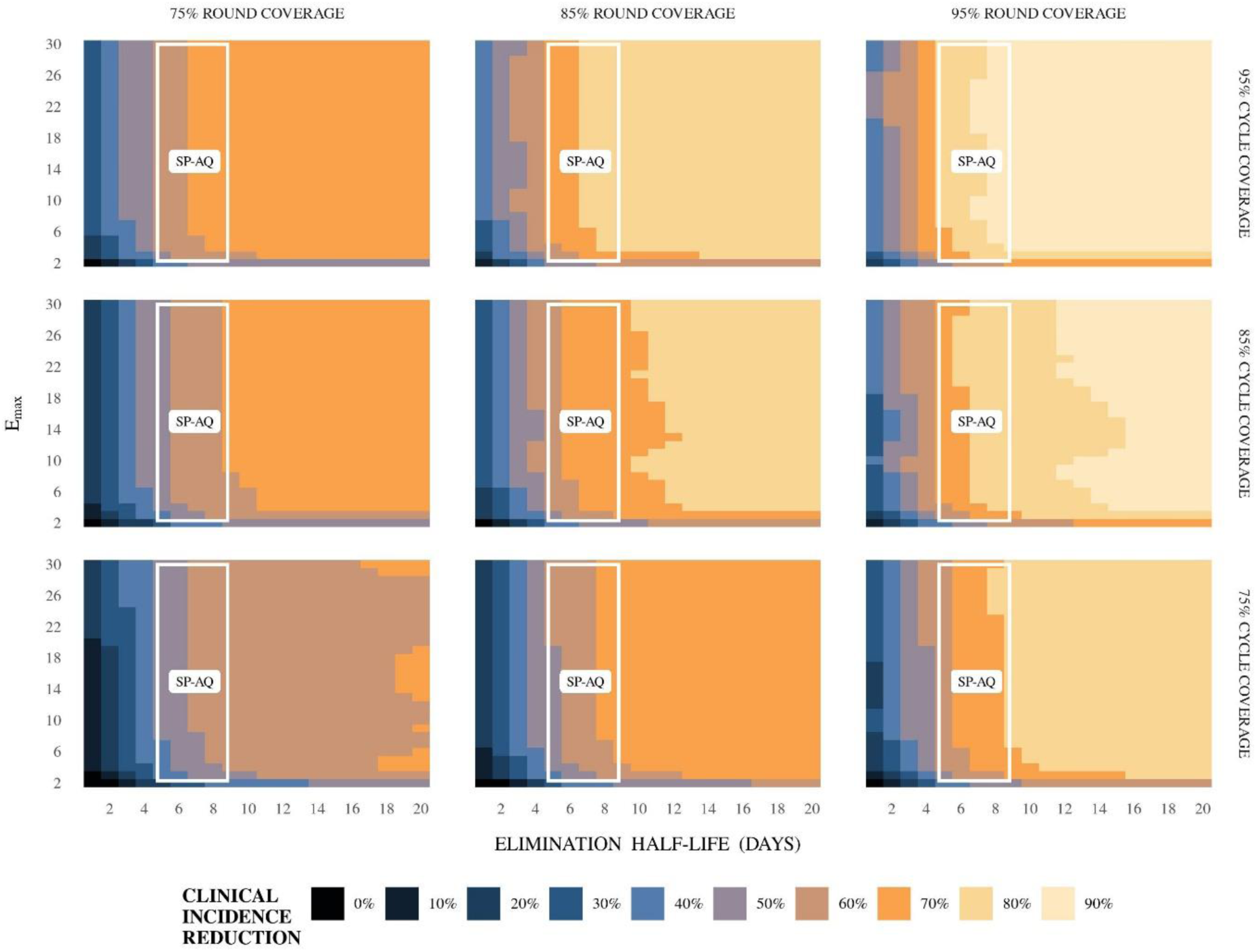
For SMC with blood stage activity only, impact of a change in coverage on the predicted relative reduction in clinical incidence (measured over a four-month intervention period) following SMC compared with a no intervention counterfactual. Each square in the grid indicates the predicted reduction (rounded to the nearest 10%) if an intervention with the given elimination half-life (horizontal axis) and maximum parasite killing rate (vertical axis) were deployed, assuming a slope of six. Variation in this figure is driven by the combined impact of stochastic uncertainty and emulator prediction error. Results are shown for children aged three and 59 months for a five-month seasonal profile with an baseline annual *Pf*PR_2-10_ of 18%, where access to first-line treatment was 50% within 14 days and where SMC was deployed four times at monthly intervals in a given year surrounding peak seasonality. Each panel represents results for a different level of SMC round coverage (75%, 85%, and 95%) and cycle coverage (75%, 85%, and 95%). The white region indicates the space of parameter profiles whose RSS (in-silico protective efficacy calculated in comparison to SP-AQ’s protective efficacy) falls within 0.1 standard deviations of the minimum (see figure 1).

**Figure A2.12:**
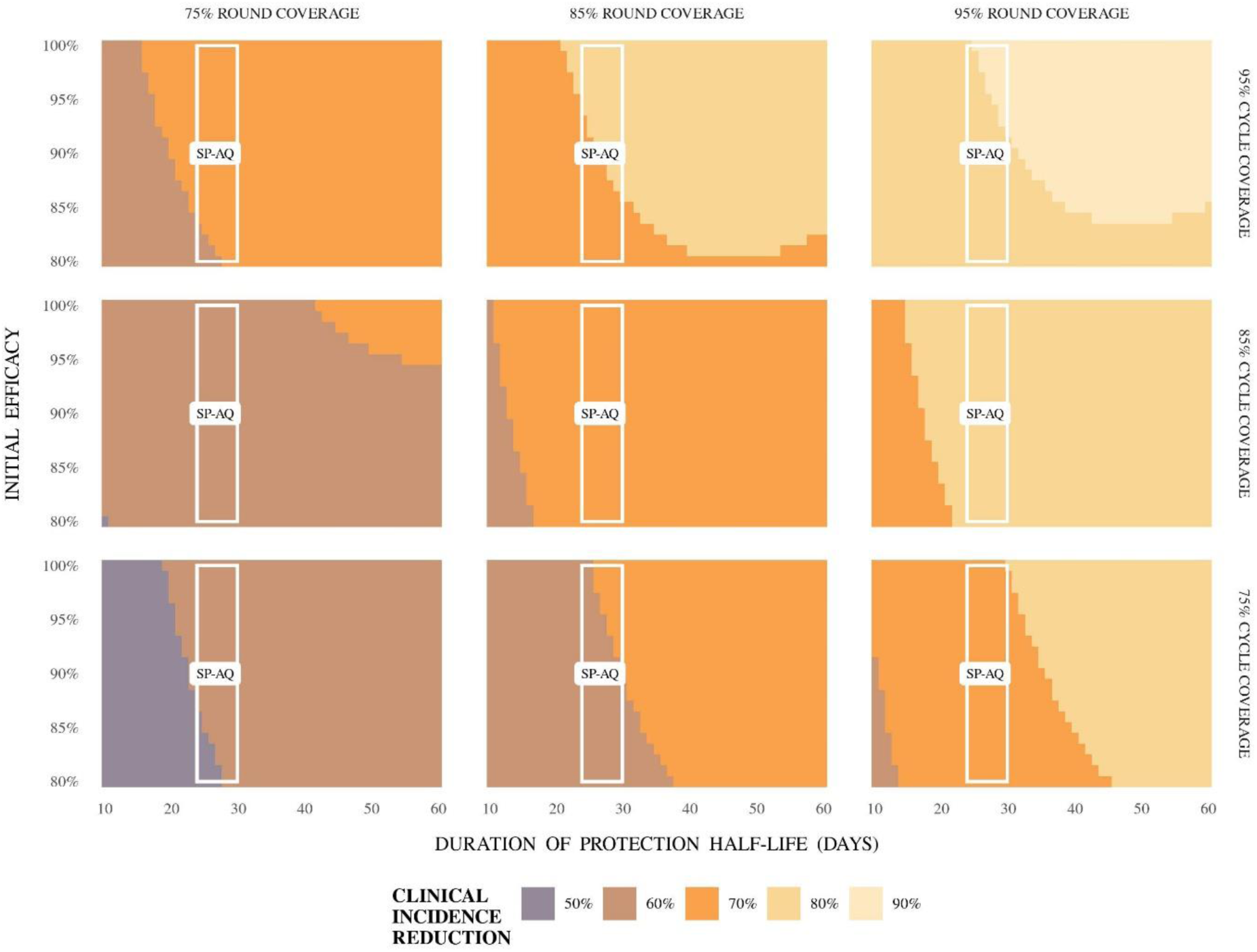
For SMC with dominant liver stage activity and initial, complete blood stage clearance, impact of a change in coverage on the predicted relative reduction in clinical incidence (measured over a four-month intervention period) following SMC compared with a no intervention counterfactual. Each square in the grid indicates the predicted reduction (rounded to the nearest 10%) if an intervention with the given duration of protection half-life (horizontal axis) and initial efficacy (vertical axis) were deployed. Variation in this figure is driven by the combination of combined impact of stochastic uncertainty and emulator prediction error. Results are shown for children aged three and 59 months for a five-month seasonal profile with an baseline annual *Pf*PR_2-10_ of 18%, where access to first-line treatment was 50% within 14 days and where SMC was deployed four times at monthly intervals in a given year surrounding peak seasonality. Each panel represents results for a different level of SMC round coverage (75%, 85%, and 95%) and cycle coverage (75%, 85%, and 95%). The white region indicates the space of parameter profiles whose RSS (in-silico protective efficacy calculatedin comparisonto SP-AQ’s protective efficacy) falls within 0.1 standard deviations of the minimum (see figure 1).

**Figure A2.13:**
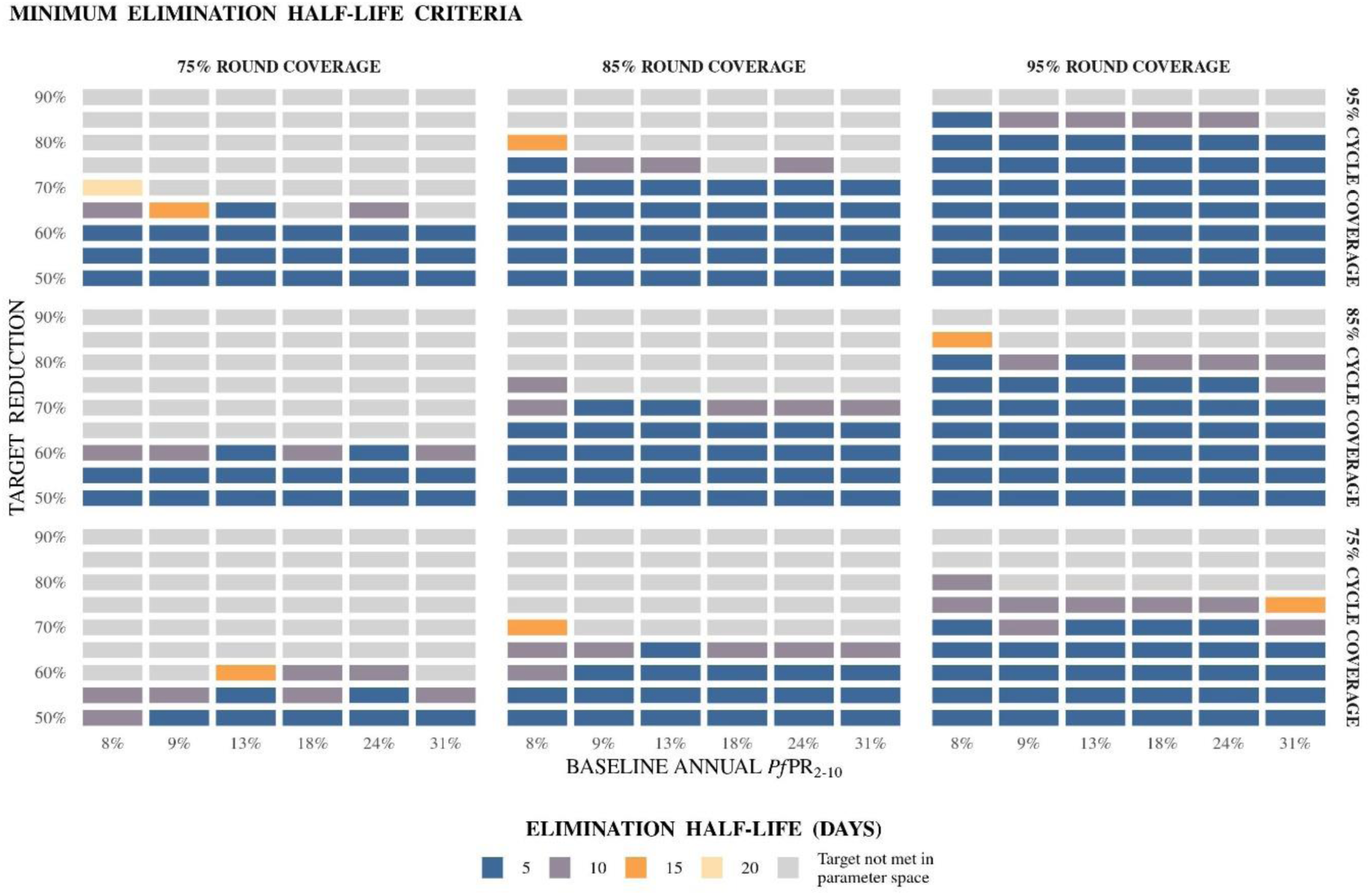
Predicted minimum elimination half-life values for SMC with dominant blood stage activity and initial, complete liver stage clearance to achieve target reductions in clinical incidence and severe disease. Summary of the predicted minimum elimination half-life criteria for SMC with dominant blood stage activity and initial, complete liver stage clearance towards achieving a target clinical incidence reduction for varying levels of SMC round and cycle coverage, shown for a five-month seasonal profile in a setting with 50% access to first-line treatment, where SMC was deployed in four monthly cycles to children three to 59 months old. Results show the estimatedminimum eliminationhalf-life witha 95% predictioninterval above the given target reductioninclinical incidence (vertical axis), compared with a no intervention counterfactual and measured across the four-month intervention period. Minimum criteria were calculated across E_max_ levels.

